# Accelerometry-Derived REM Sleep Behavior Disorder Score Is Associated with Incident Parkinson’s Disease in the UK Biobank

**DOI:** 10.64898/2026.07.02.26356952

**Authors:** Giorgio Ricciardiello Mejia, Andreas Brink-Kjaer, Lang Liu, Li Zhou, Katarina Gunter, Kang Hyun Ryu, Sajila Wickramaratne, Ankit Parekh, Ziv Gan-Or, Emmanuel During

## Abstract

Estimating Parkinson’s disease (PD) risk years before diagnosis remains an unmet need. We applied a validated machine learning classifier for REM sleep behavior disorder (RBD) detection to 7-day wrist accelerometry data in 87,975 UK Biobank participants followed for 10 years. Participants in the highest RBD risk stratum (>99^th^ percentile) had an approximately fivefold increased hazard of incident PD compared with the lowest-risk group (0–90^th^ percentile), with a dose-dependent relationship across the full score distribution. Among non-converters, higher RBD-classifier scores were associated with baseline cognitive deficits and greater burden of autonomic and psychiatric prodromal features. The association with incident PD remained independent of PD polygenic risk score, while RBD-classifier score and genetic risk were synergistic. The combined high-risk group achieved a positive likelihood ratio of 7.91, approximately threefold higher than questionnaire-based RBD screening. At this High-risk threshold over the 10-year horizon, the classifier showed a sensitivity of 7.4%, specificity of 99.2%, positive predictive value of 7.5%, and negative predictive value of 99.2% for incident PD (N=50,816). These findings support wrist accelerometry as a scalable, high-specificity approach for prodromal PD risk enrichment.

## INTRODUCTION

Parkinson’s disease (PD) is the fastest-growing neurological disorder worldwide, with recent estimates suggesting that more than 8.5–11 million people are currently living with the disease globally ^1–5^. Neuropathologically, the vast majority of PD is characterized by progressive alpha-synuclein aggregation that begins years to decades before the onset of classic motor symptoms ^6^. This prolonged prodromal interval represents a critical therapeutic window during which high-risk individuals could potentially be identified prior to irreversible neurodegeneration and clinical diagnosis.

Idiopathic REM sleep behavior disorder (iRBD)––characterized by failure of physiological REM sleep atonia and dream-enactment behavior––affects about 1% of middle-aged and older adults and is among the most specific prodromal markers of α-synucleinopathies ^7^. RBD affects approximately one-quarter of people with newly diagnosed PD, with prevalence increasing over the course of the disease ^8–10^. Longitudinal studies of polysomnography-confirmed iRBD demonstrate annual phenoconversion rates of approximately 6.3%, with most participants eventually developing PD or dementia with Lewy bodies, and less commonly multiple system atrophy ^11,12^. Accordingly, the Movement Disorder Society (MDS) prodromal criteria assign polysomnography-confirmed RBD a likelihood ratio of 130 for future PD––higher than that of any other single prodromal marker ^13,14^– –compared with 2.8 for questionnaire-based RBD assessment, reflecting the challenge of distinguishing true RBD from mimics without objective testing. Yet polysomnography remains costly, labor-intensive, and impractical for population-scale screening.

Recent advances in wearable sensing technology and machine learning (ML) have enabled the development of accelerometry-based approaches for RBD detection ^15–18^. Wrist-worn wearables can capture abnormal nocturnal motor activity associated with REM sleep without atonia and have demonstrated sensitivity exceeding 90% against polysomnography in clinical cohorts ^10,15,19,20^. As wearable devices become increasingly widespread, accelerometer-derived physiological biomarkers may provide a scalable approach for identifying participants at elevated neurodegenerative risk outside specialized sleep laboratories. Whether such classifiers generalize beyond the controlled clinical settings in which they are trained to the noisier, lower-prevalence signal environment of the general population remains unknown. Distinct from prior UK Biobank studies that derive and model activity-based features for PD prediction directly within the cohort, this study asks whether a classifier trained exclusively on clinical, polysomnography-confirmed RBD status, without training on PD outcomes or retraining in the UK Biobank, can prospectively stratify incident PD risk.

The UK Biobank (UKBB) accelerometer substudy, comprising 106,053 participants who wore an Axivity AX3 (Axivity Ltd, Newcastle, United Kingdom) device for 7 days ^21^, provides a unique opportunity to address this directly. Four key questions guided this study: (1) whether an accelerometry-derived RBD-classifier score is associated with incident PD in a large population-based cohort; (2) whether the prodromal non-motor and cognitive profile of participants stratified by RBD-classifier score is consistent with early synucleinopathy; (3) whether this signal remains informative independently of inherited genetic susceptibility––including polygenic risk scores and *GBA1* carrier status––thereby suggesting that accelerometry-derived RBD signature captures prodromal risk beyond that identified by genomics alone ^13,14^; and (4) whether accelerometry-derived RBD scoring possesses sufficient discriminatory utility to function as a scalable enrichment biomarker for population-level PD risk stratification.

Based on the established association between polysomnography-confirmed RBD and PD phenoconversion ^11–14,22,23^, we formulated a priori hypotheses for each aim: (1) higher accelerometry-derived RBD-classifier scores would be associated with increased incident PD risk; (2) participants with high RBD-classifier scores would show a greater burden of non-motor prodromal and cognitive features consistent with early synucleinopathy ^13,14^; (3) the RBD-classifier score would remain associated with incident PD after adjustment for polygenic risk and GBA1 carrier status, reflecting a distinct, non-genetic dimension of risk; and (4) accelerometry-derived RBD scoring would demonstrate sufficient discriminative performance to support its use as a population-level enrichment biomarker, with an anticipated trade-off favoring specificity over sensitivity given the rarity of the outcome in an unselected population.

To this end, we applied an externally trained ^24^ and validated ^25^ accelerometry-derived RBD classifier, without retraining, to 87,975 UK Biobank participants with valid accelerometry data, examining incident PD over a median follow-up of 10 (1.4) years using longitudinal survival analyses (**Figure 1**).

**Figure 1.**
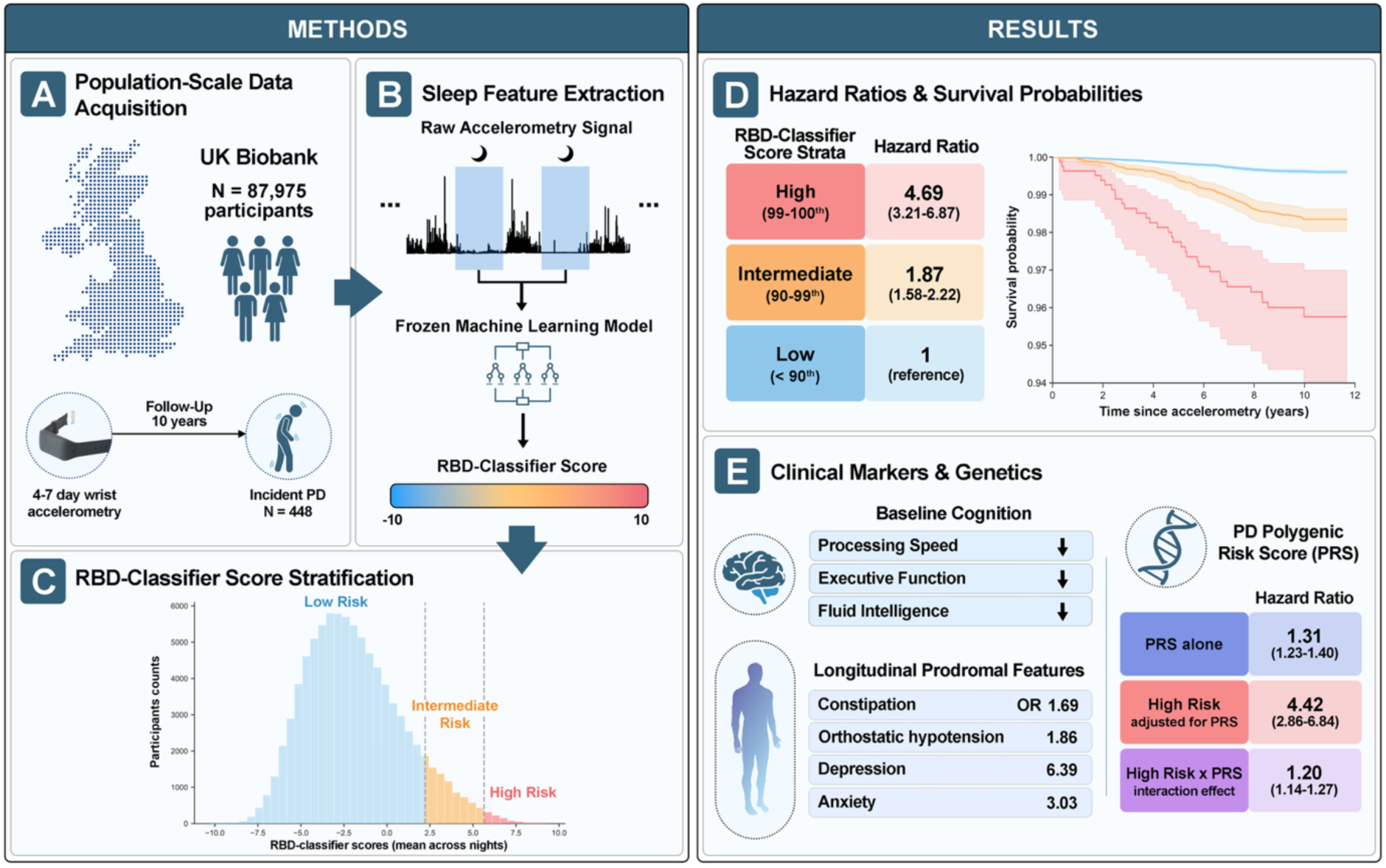
Study overview. **(A) Population-scale data acquisition.** Wrist-worn accelerometry recordings from 87,975 UK Biobank participants were utilized and linked to the longitudinal health records, yielding 448 incident Parkinson’s disease (PD) cases over approximately 10 years of follow-up. **(B) Sleep feature extraction**. Nocturnal accelerometry signals were processed using a previously trained and validated (42 cases and 42 controls) boosted decision tree classifier to generate a continuous accelerometry-derived RBD risk score. **(C) RBD-Classifier score stratification**. Participants were categorized into Low (0–90th percentile), Intermediate (90–99th percentile), and High (99–100th percentile) RBD-classifier score groups based on the distribution of risk scores. **(D) Hazard ratios and survival probabilities**. Increasing RBD-classifier scores were associated with progressively higher risk of incident PD, with participants in the highest-risk stratum exhibiting a 4.69-fold (95% CI 3.21–6.87) higher hazard of PD compared with the low-risk group. Kaplan-Meier survival curves demonstrated early and sustained separation across risk strata. **(E) Clinical markers and genetics.** Higher RBD-classifier scores were associated with enrichment of cognitive deficits and prodromal features, while the association with incident PD remained largely independent of polygenic risk. *Figure elements adapted from BioRender.com. Actigraph image courtesy of Axivity Ltd (www.axivity.com).

## RESULTS

### Cohort characteristics

The analytical cohort included 87,975 participants with valid wrist accelerometry recordings and no prevalent PD at baseline (**Supplementary Table 1**), among whom 448 incident PD cases occurred over a median follow-up of 10 (1.4) years. Accelerometry-derived RBD-classifier scores were derived from 6.59 ± 0.86 nights. Participants were stratified into percentile-based accelerometry-derived RBD risk groups: Low (0–90th percentile), Intermediate (90–99th percentile), and High (99– 100th percentile) (**Supplementary Table 2**).

Baseline characteristics differed across RBD-classifier score strata (**Table 1**). Participants in the High-score group were descriptively older (59.6 vs. 55.9 years), more often male (75% vs. 41%), had higher BMI, and showed higher observed prevalence of depression, constipation, anxiety, orthostatic hypotension, and erectile dysfunction than participants in the Low-score group. The High-score group also showed poorer performance on several baseline cognitive measures, including reaction time, numeric memory, and Trail-Making Test A/B. Statistically significant test across the three strata were significant for these variables (Kruskal–Wallis and chi-squared tests); pairwise post-hoc comparisons between individual strata were not performed.

**Table 1.**
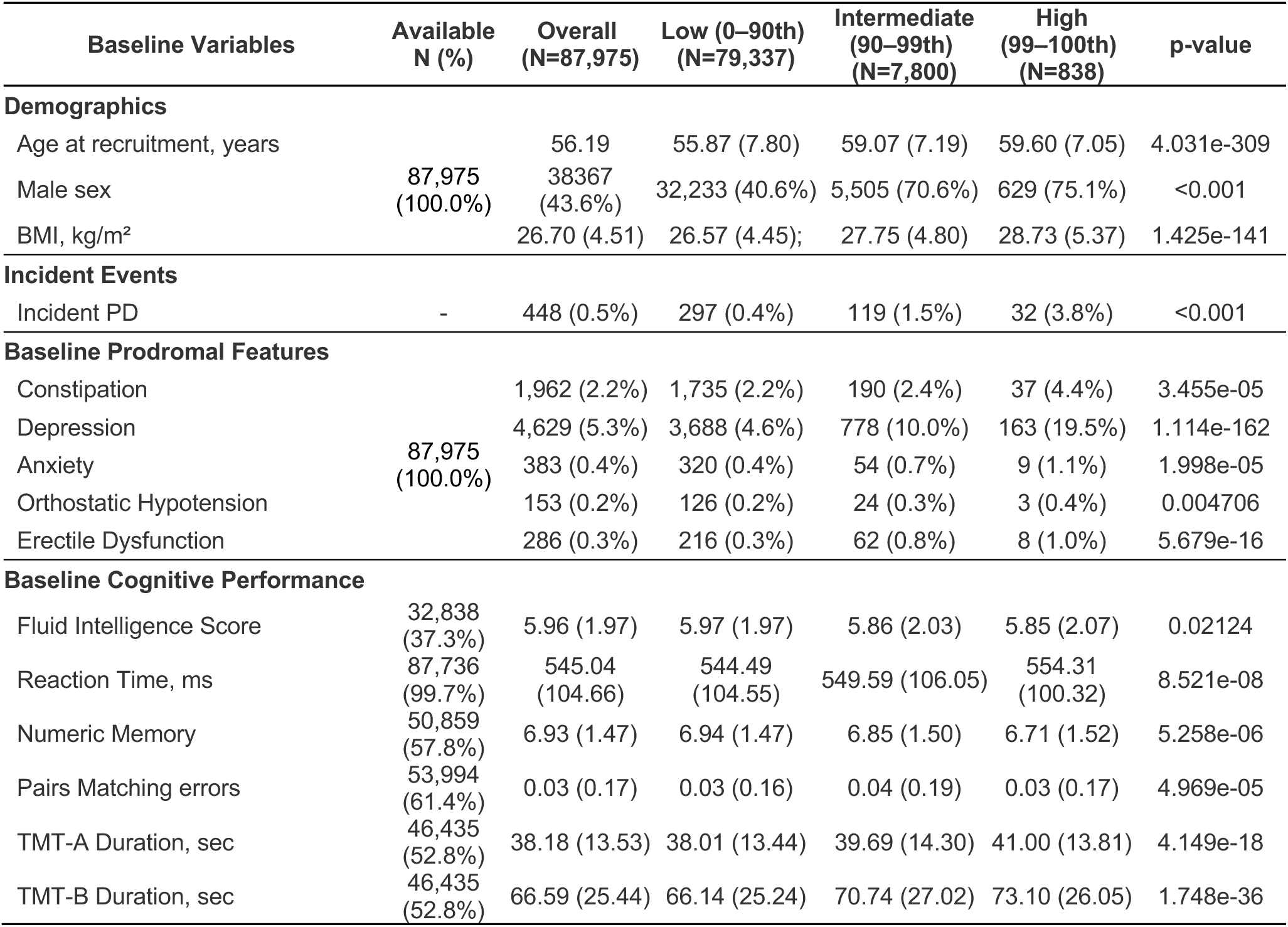
Baseline Characteristics, Prodromal Features, and Cognitive Performance by Wearable-Derived RBD Risk Stratum. Data presented as mean (SD) for continuous variables and n (%) for categorical variables. P-values reflect omnibus comparisons across the three strata using Kruskal–Wallis tests for continuous variables and chi-squared tests for categorical variables. No pairwise post-hoc comparisons between individual strata were performed; between-stratum differences described in the text are therefore descriptive. Cognitive performance was not completed by all participants at baseline.

| **Baseline Variables** | **Available N (%)** | **Overall (N=87,975)** | **Low (0–90th) (N=79,337)** | **Intermediate (90–99th) (N=7,800)** | **High (99–100th) (N=838)** | **p-value** |
|---|---|---|---|---|---|---|
| **Demographics** |  |  |  |  |  |  |
| Age at recruitment, years | 87,975 (100.0%) | 56.19 | 55.87 (7.80) | 59.07 (7.19) | 59.60 (7.05) | 4.031e-309 |
| Male sex |  | 38367 (43.6%) | 32,233 (40.6%) | 5,505 (70.6%) | 629 (75.1%) | <0.001 |
| BMI, kg/m² |  | 26.70 (4.51) | 26.57 (4.45); | 27.75 (4.80) | 28.73 (5.37) | 1.425e-141 |
| **Incident Events** |  |  |  |  |  |  |
| Incident PD | - | 448 (0.5%) | 297 (0.4%) | 119 (1.5%) | 32 (3.8%) | <0.001 |
| **Baseline Prodromal Features** |  |  |  |  |  |  |
| Constipation | 87,975 (100.0%) | 1,962 (2.2%) | 1,735 (2.2%) | 190 (2.4%) | 37 (4.4%) | 3.455e-05 |
| Depression |  | 4,629 (5.3%) | 3,688 (4.6%) | 778 (10.0%) | 163 (19.5%) | 1.114e-162 |
| Anxiety |  | 383 (0.4%) | 320 (0.4%) | 54 (0.7%) | 9 (1.1%) | 1.998e-05 |
| Orthostatic Hypotension |  | 153 (0.2%) | 126 (0.2%) | 24 (0.3%) | 3 (0.4%) | 0.004706 |
| Erectile Dysfunction |  | 286 (0.3%) | 216 (0.3%) | 62 (0.8%) | 8 (1.0%) | 5.679e-16 |
| **Baseline Cognitive Performance** |  |  |  |  |  |  |
| Fluid Intelligence Score | 32,838 (37.3%) | 5.96 (1.97) | 5.97 (1.97) | 5.86 (2.03) | 5.85 (2.07) | 0.02124 |
| Reaction Time, ms | 87,736 (99.7%) | 545.04 (104.66) | 544.49 (104.55) | 549.59 (106.05) | 554.31 (100.32) | 8.521e-08 |
| Numeric Memory | 50,859 (57.8%) | 6.93 (1.47) | 6.94 (1.47) | 6.85 (1.50) | 6.71 (1.52) | 5.258e-06 |
| Pairs Matching errors | 53,994 (61.4%) | 0.03 (0.17) | 0.03 (0.16) | 0.04 (0.19) | 0.03 (0.17) | 4.969e-05 |
| TMT-A Duration, sec | 46,435 (52.8%) | 38.18 (13.53) | 38.01 (13.44) | 39.69 (14.30) | 41.00 (13.81) | 4.149e-18 |
| TMT-B Duration, sec | 46,435 (52.8%) | 66.59 (25.44) | 66.14 (25.24) | 70.74 (27.02) | 73.10 (26.05) | 1.748e-36 |

### RBD-classifier Score Stratifies Incident Parkinson’s Disease

Accelerometry-derived RBD-classifier scores strongly stratify future PD risk across the general population cohort. PD incidence increased progressively across RBD strata, rising from 0.4% in the Low-RBD risk group to 1.5% in the Intermediate-risk group and 3.8% in the High-risk group.

Compared with the Low-risk reference group, participants in the High-risk stratum exhibited a markedly elevated hazard of incident PD (HR 4.69, 95% CI 3.21–6.87; p = 1.80 × 10^−15^), while the Intermediate-RBD group demonstrated a more modest but still significant increase (HR 1.87, 95% CI 1.58–2.22; p = 6.81 × 10^−13^) (**Supplementary Table 3**). Similar associations were observed when modeling the RBD-classifier score continuously, with each standard deviation increase associated with a 28% higher PD hazard (HR 1.28, 95% CI 1.21–1.35; p = 8.8 × 10^−20^; Harrell’s C-index 0.784), a measurement of model discrimination ranging from 0.5 (not better than chance) to 1 (perfect discrimination).

### PD Risk Concentrates at Extreme RBD-Classifier Scores

Restricted cubic spline modeling demonstrated a highly significant non-linear association between RBD-classifier score and incident PD risk (non-linearity p = 5.3 × 10^−13^). Rather than increasing uniformly across the score distribution, PD hazard accelerated sharply at the extreme upper tail (**Figure 2**).

**Figure 2.**
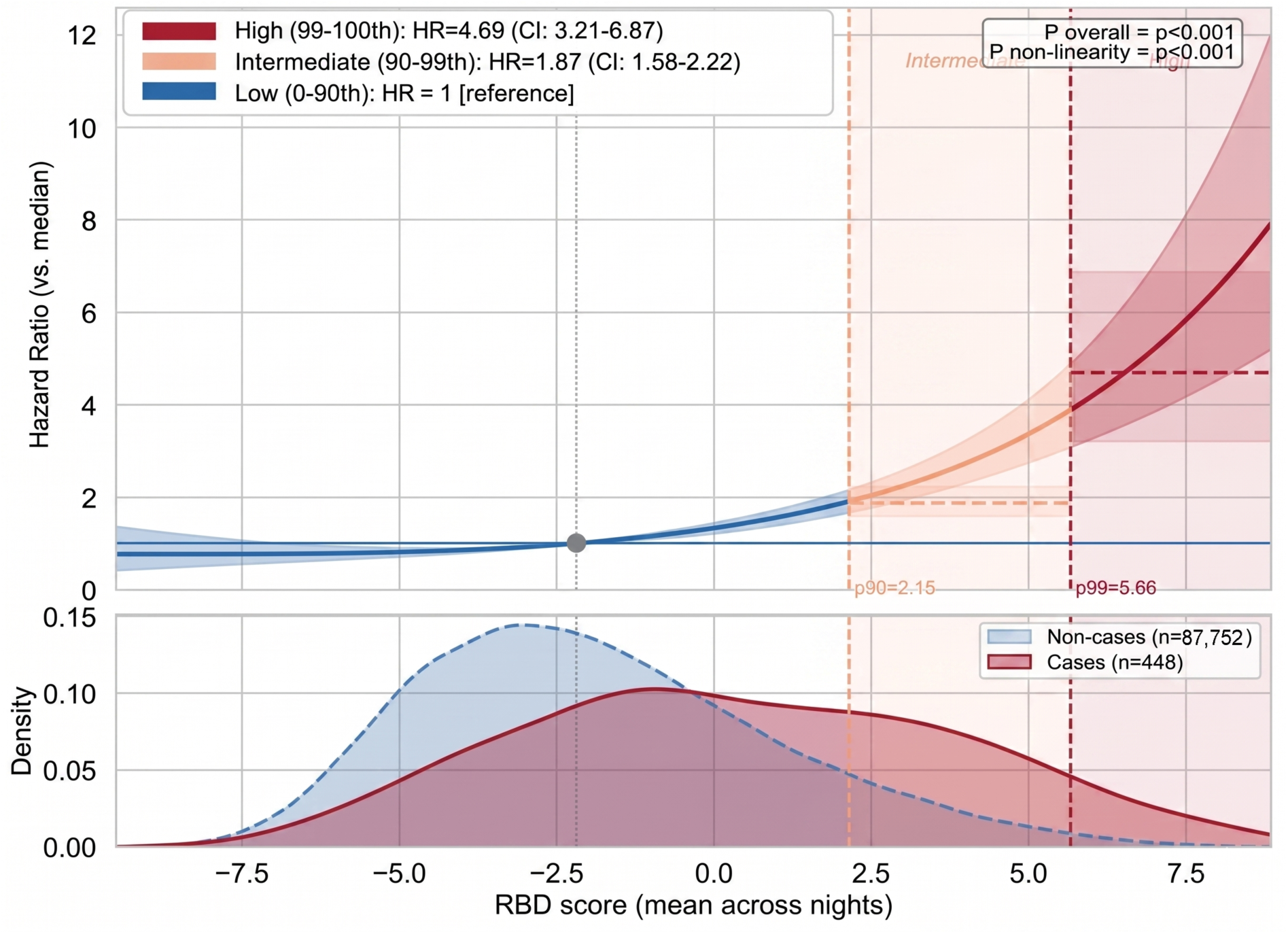
Continuous dose–response relationship between RBD-classifier score and incident PD modeled using restricted cubic spline Cox regression. Vertical dashed lines indicate percentile thresholds corresponding to categorical risk groups. Models were adjusted for age, sex, BMI, smoking, and alcohol use.

Consistent with this pattern, participants in the High-risk group who later developed PD were diagnosed with PD approximately one year earlier than participants in the lower-risk strata (median time-to-diagnosis 4.66 vs 5.62 years; p = 6.6 × 10^−8^). This temporal compression suggests that the highest accelerometry-derived RBD-classifier scores capture a distinct prodromal subgroup closer to phenoconversion rather than simply reflecting increasing nonspecific sleep disturbance.

### RBD-Classifier Track Prodromal Features in Non-Converters

Among non-converters, higher RBD-classifier scores were associated with greater prevalence of prodromal features at across the observational period (**Table 2**). This relationship was most pronounced for depression (Low: 5.9% vs. High: 21.4%) and constipation (Low: 5.3% vs. High: 10.2%), and remained significant across all prodromal markers except anosmia, for which events were too sparse for meaningful analysis (N=11). Baseline and follow-up prodromal markers are presented separately in **Supplementary Table 4**.

**Table 2.**
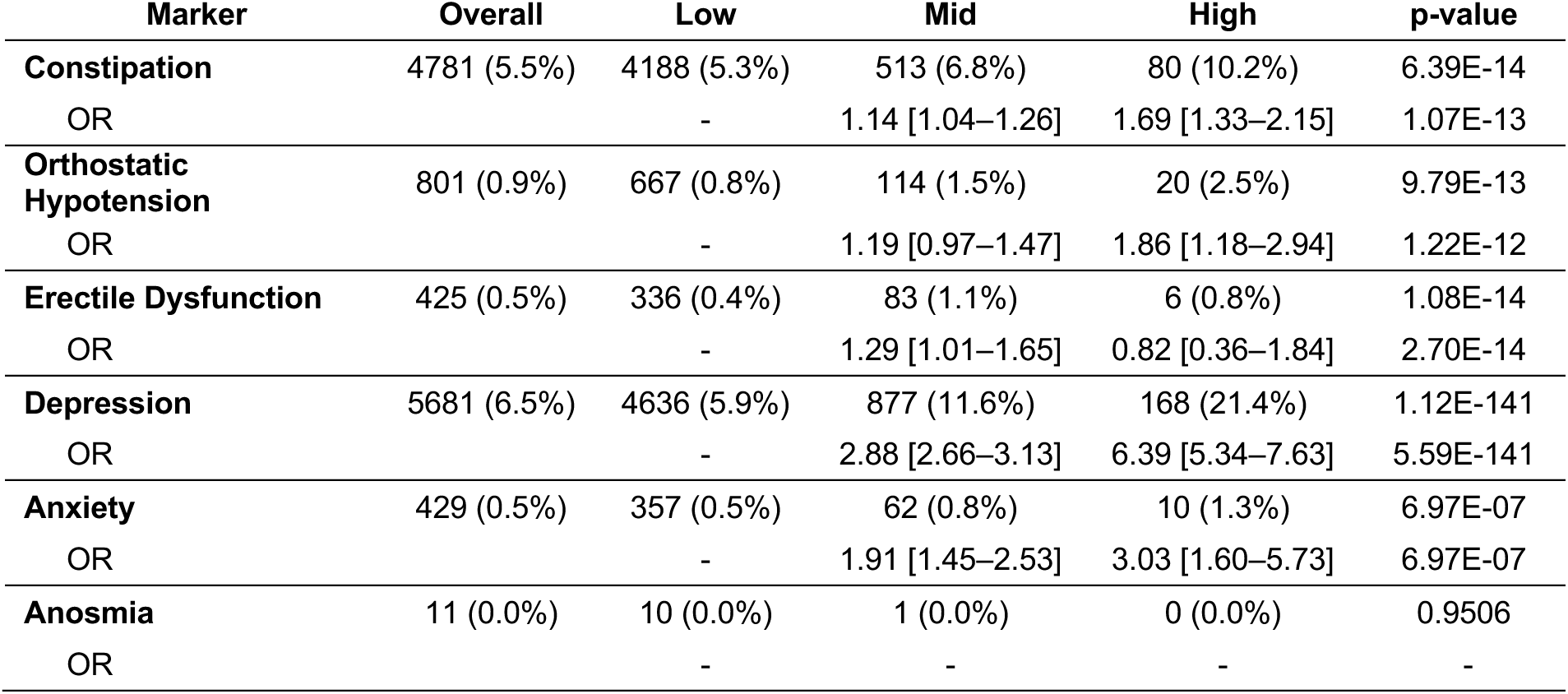
Distribution of prodromal features by actigraphy-derived RBD risk group among non-converters. Values represent the number and percentage of participants with each prodromal feature at any assessment (baseline or subsequent visit) among participants who were not diagnosed with PD or any dementia (N = 86,973). Participants were stratified into Low (0–90th percentile), Intermediate (90–99th percentile), and High (99–100th percentile) groups according to accelerometry-derived RBD-classifier score. Presence of a prodromal feature was defined as occurrence at either baseline or follow-up. P-values were derived from χ^2^ tests comparing prevalence across RBD strata. Odds ratios (OR) are presented with 95% confidence intervals (CI) from logistic regression models adjusted for age at recruitment, sex, and total follow-up time.

| Marker | Overall | Low | Mid | High | p-value |
|---|---|---|---|---|---|
| **Constipation** | 4781 (5.5%) | 4188 (5.3%) | 513 (6.8%) | 80 (10.2%) | 6.39E-14 |
| OR |  | - | 1.14 [1.04–1.26] | 1.69 [1.33–2.15] | 1.07E-13 |
| **Orthostatic Hypotension** | 801 (0.9%) | 667 (0.8%) | 114 (1.5%) | 20 (2.5%) | 9.79E-13 |
| OR |  | - | 1.19 [0.97–1.47] | 1.86 [1.18–2.94] | 1.22E-12 |
| **Erectile Dysfunction** | 425 (0.5%) | 336 (0.4%) | 83 (1.1%) | 6 (0.8%) | 1.08E-14 |
| OR |  | - | 1.29 [1.01–1.65] | 0.82 [0.36–1.84] | 2.70E-14 |
| **Depression** | 5681 (6.5%) | 4636 (5.9%) | 877 (11.6%) | 168 (21.4%) | 1.12E-141 |
| OR |  | - | 2.88 [2.66–3.13] | 6.39 [5.34–7.63] | 5.59E-141 |
| **Anxiety** | 429 (0.5%) | 357 (0.5%) | 62 (0.8%) | 10 (1.3%) | 6.97E-07 |
| OR |  | - | 1.91 [1.45–2.53] | 3.03 [1.60–5.73] | 6.97E-07 |
| **Anosmia** | 11 (0.0%) | 10 (0.0%) | 1 (0.0%) | 0 (0.0%) | 0.9506 |
| OR |  | - | - | - | - |

After adjusting for age, sex, and follow-up time, the association between accelerometry-derived RBD risk and prodromal features remained robust across markers (**Table 2; Supplementary Table 5**). The effect was strongest for depression: High-risk participants had 6.4-fold greater odds of depression compared with Low-risk participants (OR 6.39 [95% CI 5.34–7.63]), and Intermediate-risk participants had nearly threefold greater odds (OR 2.88 [95% CI 2.66–3.13]). Anxiety showed a similar dose-response gradient (Intermediate: OR 1.91 [1.45–2.53]; High: OR 3.03 [1.60–5.73]). Constipation odds increased monotonically across strata (Intermediate: OR 1.14 [1.04–1.26]; High: OR 1.69 [1.33–2.15]). Orthostatic hypotension was significantly elevated only in the High-risk group (OR 1.86 [1.18–2.94], p=0.008), with no significant effect in the Intermediate stratum (OR 1.19 [0.97–1.47], p=0.09).

### RBD-Classifier Show Preferential Association With PD

To compare associations across neurodegenerative outcomes, we examined associations between RBD-classifier score and alternative neurodegenerative outcomes (**Supplementary Table 6**). In contrast to the strong PD association, relationships with Alzheimer’s disease (HR 1.51 [95% CI 0.87–2.63]; p = 0.145) and vascular dementia (HR 1.52 [95% CI 0.81–2.88]; p = 0.194) were substantially attenuated and did not reach statistical significance. These findings argue against the observed signal simply reflecting generalized frailty, poor sleep quality, or non-specific neurological vulnerability, and instead support its preferential alignment with prodromal α-synucleinopathy.

### RBD-Classifier Physiology Captures PD Risk Beyond Genetics

In the genetic subcohort (N = 66,524; 359 incident PD cases), adjusting for PD polygenic risk score (PD-PRS) and ancestry principal components had minimal impact on the RBD association (**Figure 3**). The continuous RBD effect remained strongly associated with incident PD after PRS adjustment (HR 1.27 per SD [95% CI 1.20–1.35]; p = 3.5 × 10^−15^), while High-risk participants retained approximately 4-fold elevated hazard (HR 4.42 [95% CI 2.86–6.84]).

**Figure 3.**
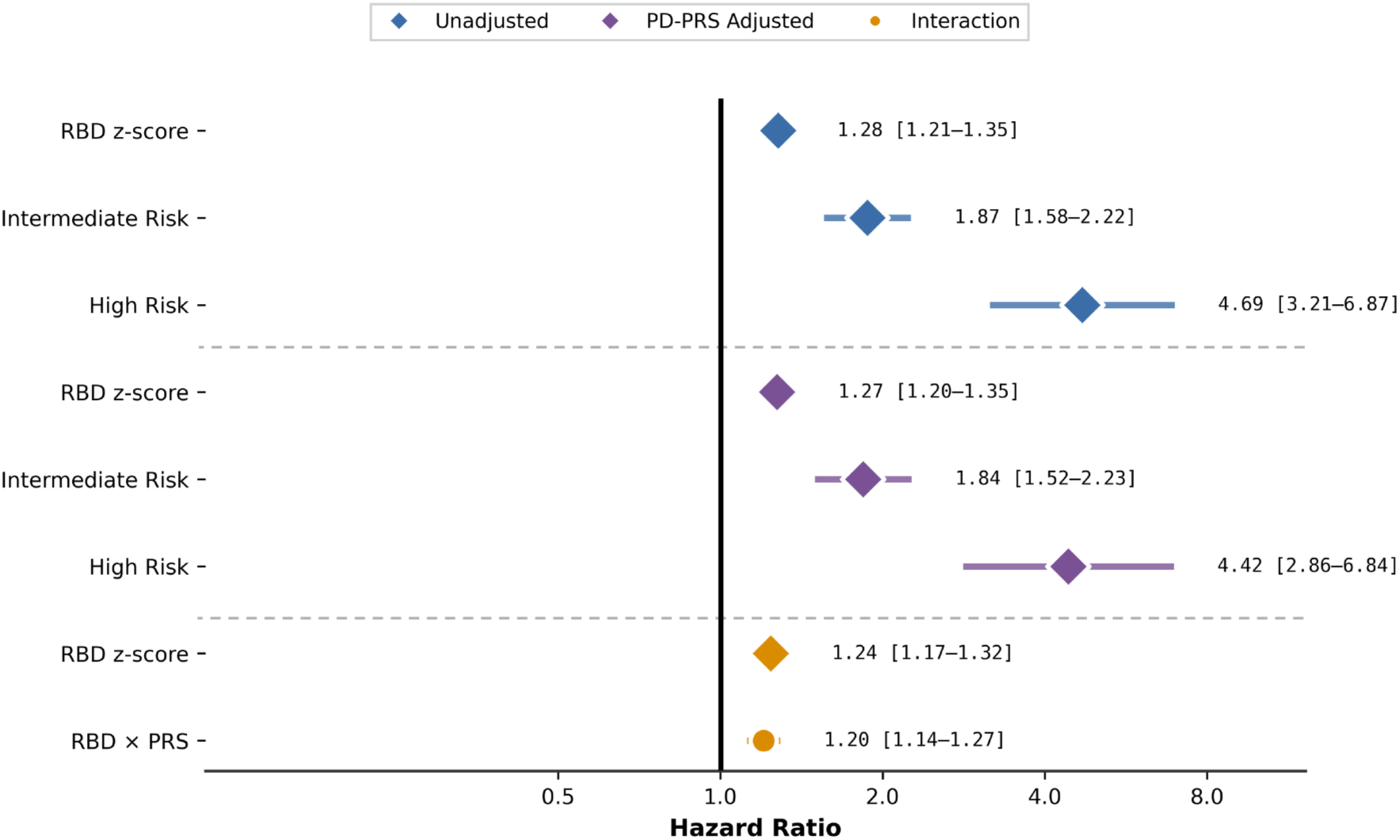
Association between accelerometry-derived RBD risk score, polygenic risk, and incident Parkinson’s disease. Forest plot showing hazard ratios for incident Parkinson’s disease associated with accelerometry-derived RBD measures before and after adjustment for Parkinson’s disease polygenic risk score (PD-PRS). The association between accelerometry-derived RBD physiology and incident PD remained largely unchanged after PRS adjustment, with the high-RBD risk group retaining approximately fourfold elevated hazard (unadjusted HR 4.69 [3.21–6.87] vs. PRS-adjusted HR 4.42 [2.86–6.84]). Continuous interaction modeling demonstrated significant amplification of PD risk in participants with both elevated RBD-classifier scores and higher genetic susceptibility (RBD × PRS interaction HR 1.20 [1.14–1.27]). These findings support that accelerometry-derived RBD physiology captures a dimension of prodromal PD risk that is complementary to inherited genetic susceptibility.

PD-PRS independently predicted PD risk with a HR of 1.31 per SD, 95% CI 1.28–1.35 (*μ* =0.021; SD=0.98). When PD-PRS is included with the RBD risk groups in the Cox regression model, the PD-PRS HR is 1.36 per SD, 95% CI 1.28–1.45. At the interaction level, participants with both elevated RBD-classifier scores and higher PD-PRS exhibited significantly increased risk (RBD × PD-PRS interaction HR 1.20 [95% CI 1.14–1.27]; p = 3.2 × 10^−10^; C-index 0.83), indicating that accelerometry-derived physiology and genetic risk capture complementary dimensions of vulnerability (**Supplementary Tables 7-9**).

Exploratory *GBA1* analyses were underpowered (166 carriers; 4 incident PD cases) but demonstrated a directional interaction between continuous RBD-classifier score and carrier status (HR 2.10 [95% CI 0.99–4.44]; p = 0.053; **Supplementary Table 10**).

### RBD-Classifier Scores Add Beyond Clinical Prodromal Features

Baseline prodromal markers alone showed limited predictive value for incident PD, with no marker remaining significant after multiple-comparison correction in prodromal-only models (**Supplementary Table 11**). In additive models, the association between accelerometry-derived RBD-classifier score and PD remained largely unchanged after adjustment for individual baseline prodromal features, with High-risk participants consistently demonstrating approximately 4.6–6.1-fold elevated PD hazard (**Supplementary Table 12**).

Interaction analyses showed no consistent multiplicative or additive synergy between RBD-classifier score and baseline prodromal markers (**Supplementary Table 13**), and absolute-risk analyses demonstrated minimal additional enrichment beyond RBD prediction alone (**Supplementary Tables 14-15**). Similarly, executive function as measured by the TMT-B/A ratio was not independently associated with PD risk, while the RBD association remained robust (**Supplementary Table 16**).

### Robustness and Model Discrimination

The association between accelerometry-derived RBD-classifier score and incident PD remained robust across multiple sensitivity analyses. Competing-risk adjustment using the Aalen–Johansen cumulative incidence function (CIF) demonstrated negligible bias relative to Kaplan–Meier estimates. CIF and Kaplan–Meier curves were virtually identical across all RBD strata, with absolute differences of 0.0 percentage points at both 5 and 10 years. At 10 years, cumulative PD incidence was 4.24%, 1.65%, and 0.38% in the High-, Intermediate-, and Low-risk groups, respectively (**Supplementary Table 17**).

Importantly, the HR association between incident PD and the RBD High-risk group remained robust across alternative percentile thresholds, including top 5%, top 10%, and top 15% cutoffs (**Supplementary Table 18**), indicating that the observed risk enrichment was not dependent on a single arbitrary threshold definition. Predictive performance for the 90^th^, 95^th^, and 99^th^ percentiles were evaluated at 5- and 10-year time horizons, with “screen positive” defined by the percentile threshold and “true positive” defined as an incident PD event within the specified horizon; participants censored before the horizon without an event were excluded, as their eventual status could not be ascertained (Methods), with increasing specificity at the expense of sensitivity. At the 90th percentile threshold, 10-year sensitivity was 34% and specificity 91%, identifying approximately one-third of future PD cases while maintaining high specificity (N=50,816; **Supplementary Table 19**). Increasing threshold stringency progressively enriched the High-risk subgroup, with10-year positive predictive value increased from 3% at the 90th percentile to 7% at the 99th percentile among the horizon-evaluable subsample an approximately 6- to 15-fold enrichment over the cohort’s overall 10-year PD incidence (0.51%). These horizon-restricted PPV estimates use a different denominator than the crude High-risk-group PD proportion (3.8%; **Table 1**) and the model-based 10-year cumulative incidence in the High-risk group (4.24%; **Supplementary Table 17**). Negative predictive value exceeded 99% across all thresholds, which given the cohort’s low absolute PD incidence, this largely reflects the low base rate rather than demonstrating clinical utility on its own.

To address potential reverse causality, we repeated all analyses after excluding PD cases occurring within two years of accelerometry assessment. This removed 51 early-onset cases (448 to 397 cases) but did not materially alter any associations. Hazard ratios changed by less than 0.2 absolute units, and all statistical inferences remained unchanged (**Supplementary Table 20**).

Across all model families, inclusion of the accelerometry-derived RBD-classifier score consistently improved model fit and discrimination relative to covariate-only or prodromal-marker-only models. The RBD-only model achieved a C-index of 0.782 and substantially improved model fit relative to the covariate-only model (ΔAIC = −89.9). In comparison, models containing individual prodromal markers achieved C-indices ranging from 0.757 to 0.762. Incorporation of PRS further improved predictive performance, with PRS-adjusted models achieving C-indices of 0.842–0.844. The addition of the accelerometry-derived RBD-classifier score to age and sex increased discrimination by ΔC = 0.028 (95% CI: 0.017–0.040), while subsequent inclusion of PRS yielded an additional improvement of ΔC = 0.029 (95% CI: 0.018–0.041; **Supplementary Table 21**).

### Clinical Utility and Likelihood Ratio Analysis

The continuous RBD-classifier score demonstrated modest individual-level discrimination (LR+ = 1.74; LR− = 0.51) and was significantly associated with incident PD in logistic regression: each 1-SD increase in the accelerometry-derived RBD z-score was associated with a 2.04-fold higher odds of incident PD (unadjusted OR = 2.04, 95% CI 1.87–2.21, p<0.001), attenuated to OR = 1.70 (95% CI 1.56–1.86, p<0.001) after adjustment for age, sex, and BMI. However, percentile-based stratification substantially enriched future PD risk, with the intermediate-RBD risk group showing LR = 3.04 (95% CI 2.60–3.55) and the high-RBD risk group showing LR+ = 7.91 (95% CI 5.63– 11.13). In comparison, constipation and depression showed weak discriminatory value (LR+ ≈ 1.1– 1.3), while orthostatic hypotension and erectile dysfunction demonstrated moderate associations (**Figure 4**). Bayesian post-test probability analysis used an empirically derived, age-band-specific PD risk prior estimated from this cohort (interpolated across 5-year age bands, ranging from approximately 0.1% at age 50 to 2.2% at age 70; cohort-wide 10-year incidence 0.51%) and showed that, incorporating the accelerometry-derived RBD-classifier score raised the estimated probability of PD above a 1% actionable threshold in 61.4% of participants who later developed PD, compared with 24.5% of those who did not. These findings support accelerometry-derived RBD scoring as a high-specificity enrichment biomarker for prodromal PD risk.

**Figure 4.**
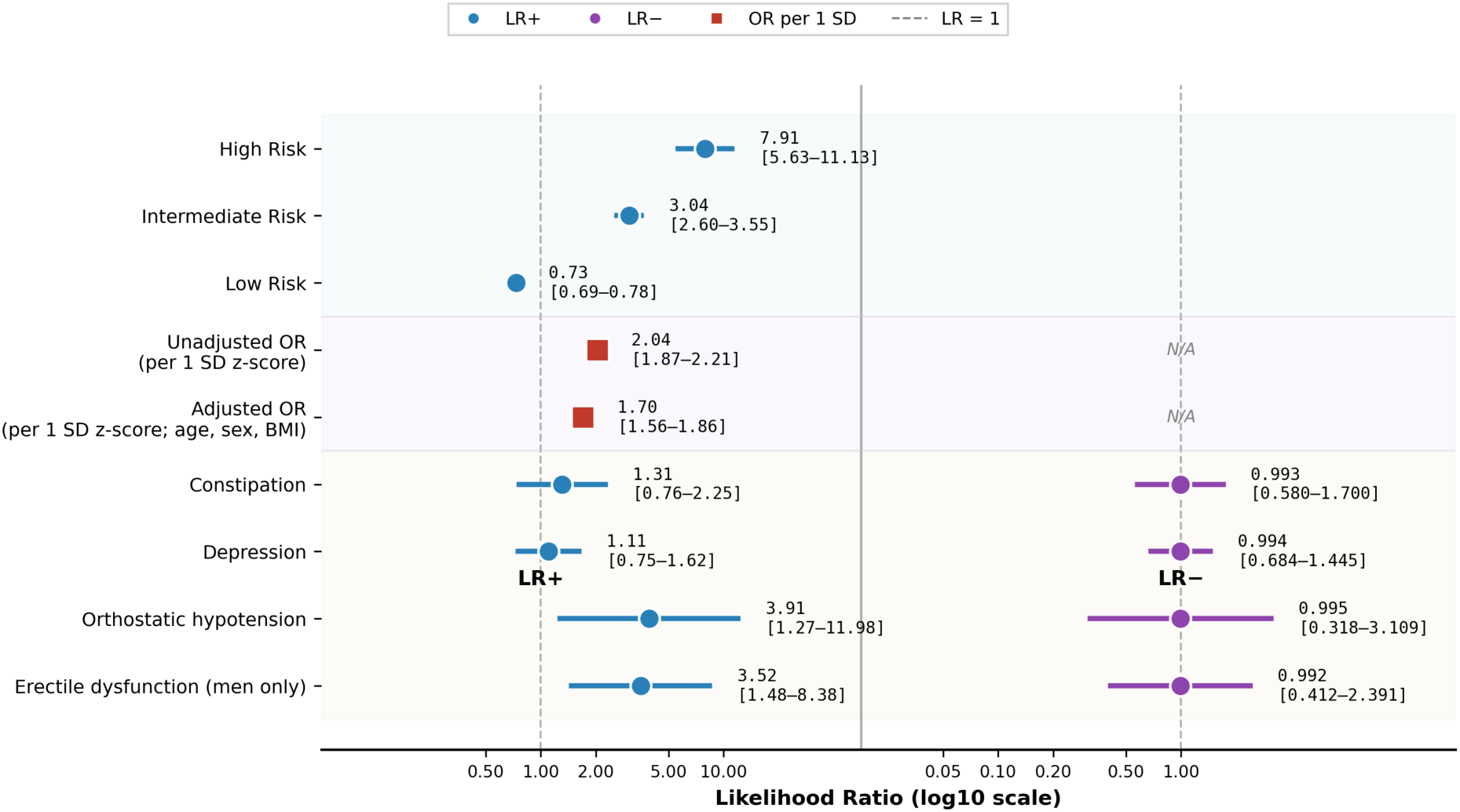
Likelihood ratios of accelerometry-derived RBD risk groups and established prodromal markers for incident Parkinson’s disease. Forest plot showing positive likelihood ratios (LR+) and negative likelihood ratios (LR−) for percentile-defined accelerometry-derived RBD risk groups and established prodromal markers. The high-RBD risk group (top 1% of the score distribution) demonstrated the strongest discriminatory performance (LR = 7.91, 95% CI 5.63–11.13), exceeding the predictive utility of traditional prodromal markers including constipation, depression, orthostatic hypotension, and erectile dysfunction. The intermediate-risk group also substantially increased future PD probability (LR = 3.04, 95% CI 2.60–3.55), whereas the low-risk group reduced disease probability (LR = 0.73, 95% CI 0.69–0.78). The continuous RBD z-score showed a significant unadjusted odds ratio of 2.04 (95% CI 1.87–2.21) per 1-SD increase, attenuating to 1.70 (95% CI 1.56–1.86) after adjustment for age, sex, and BMI, confirming that the RBD-classifier score’s association with incident PD is independent of these core confounders. Dashed vertical line indicates LR = 1 (no diagnostic discrimination).

### Cross-Sectional Prediction of Future Parkinson’s Disease

To evaluate the predictive value of accelerometry-derived RBD risk for future PD, we compared machine learning models incorporating demographics, RBD probability scores, prodromal markers, PRS, and cognitive measures. The RBD-classifier score alone demonstrated substantial discriminatory ability for identifying participants who subsequently developed PD, achieving a ROC-AUC of 0.786 when combined with demographic variables. Addition of PRS yielded the highest overall performance (ROC-AUC=0.838), whereas prodromal markers and cognitive measures provided comparatively limited incremental improvement (**Supplementary Figure 1**). Feature importance analyses further demonstrated that the accelerometry-derived RBD-classifier score was consistent among the strongest predictors across all model configurations (**Figure 5**).

**Figure 5.**
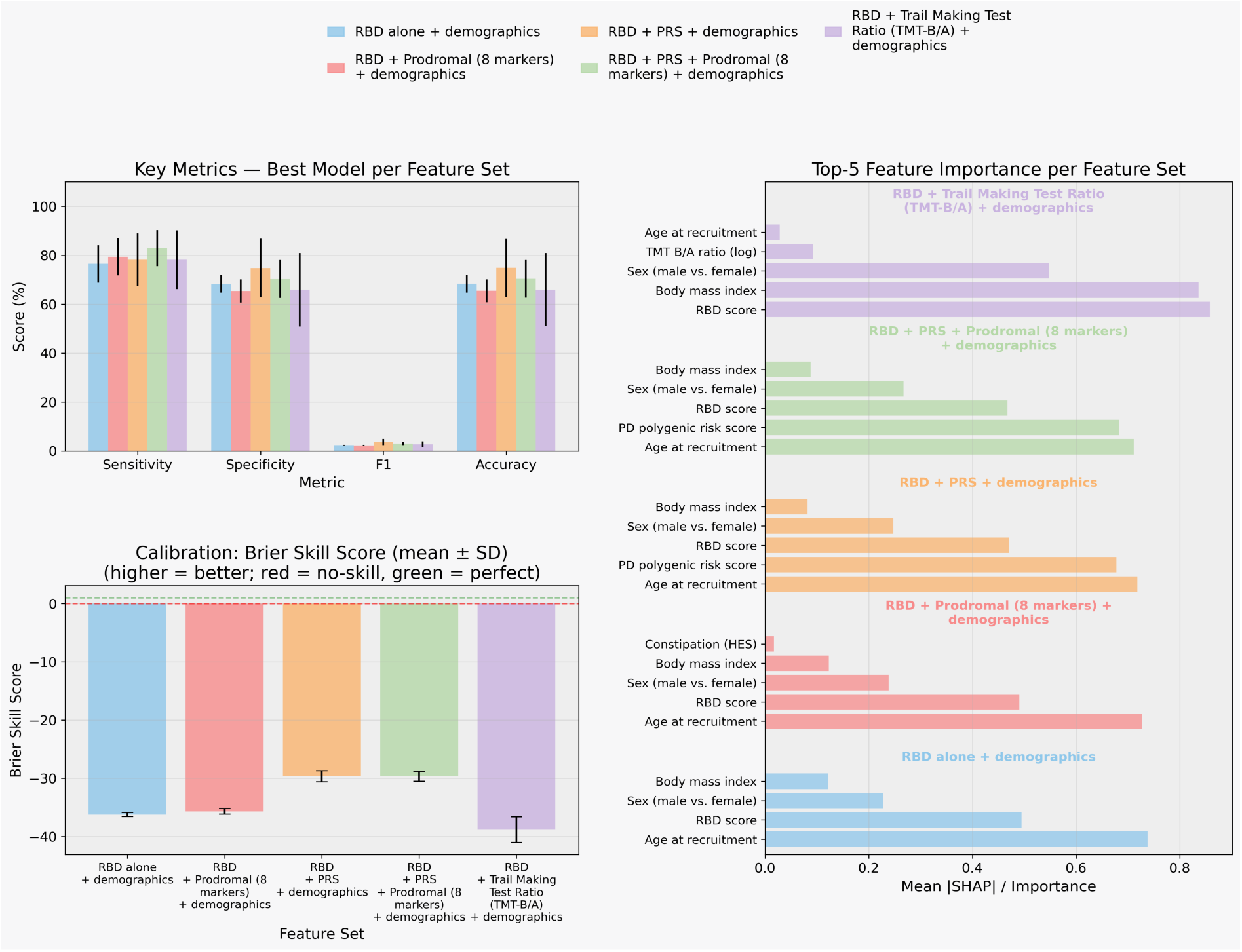
Feature-set comparison for incident Parkinson’s disease prediction. Performance metrics (sensitivity, specificity, F1-score, and accuracy), calibration (Brier Skill Score), and feature importance rankings are shown for the best-performing model within each predictor set. Predictor sets included accelerometry-derived RBD risk scores, demographic variables, prodromal markers, Parkinson’s disease polygenic risk scores (PRS), and cognitive measures. Age and RBD probability were consistently among the most influential predictors across models, while incorporation of PRS yielded the largest improvement in overall discrimination. Error bars represent standard deviations across outer cross-validation folds.

## DISCUSSION

In this prospective population-scale study of 87,975 UK Biobank participants, an accelerometry-derived RBD-classifier score strongly predicted incident PD over a median follow-up of 10 (1.4) years, with participants in the highest-risk stratum demonstrating approximately 4-fold elevated hazard and significant dose-dependent relationship across the full score distribution.

We observed a marked non-linear concentration of PD risk at the extreme upper tail of the RBD-classifier score distribution, with hazard rising sharply only among participants with the highest accelerometry-derived scores. Polysomnography-confirmed iRBD carries one of the highest known likelihood ratios for future PD ^13,14^ and prior literature has demonstrated that, within iRBD, the severity of REM sleep without atonia is a predictor of phenoconversion ^22,23^. The specificity of the RBD classifier used in this study is only 90% ^18,24,25^; therefore, RBD-classifier scores could capture a broader composite of nocturnal dysfunction beyond REM sleep, such as sleep fragmentation, periodic limb movements, and sleep-disordered breathing— all of which have been associated with adverse cognitive or neurodegenerative outcomes in longitudinal studies ^26–28^.

During the 10-year follow-up duration, RBD-classifier scores also showed a dose-dependent association with autonomic and psychiatric prodromal markers in participants who remained free of PD or dementia. Differences across risk strata were more pronounced when considering any assessment during follow-up than baseline alone, indicating a greater burden of documented prodromal features among participants with higher RBD-classifier scores. This analysis does not establish the temporal onset or progression of individual prodromal features ^29,30^. Because most cognitive assessments were conducted only at baseline, a longitudinal cognitive trajectory could not be evaluated. Nonetheless, baseline cognitive performance across strata showed graded differences in attention, processing speed, and executive function, consistent with the cognitive changes documented years before PD or dementia onset in patients with confirmed RBD. Notably, the LRs of individual prodromal markers were largely concordant with MDS benchmark values ^13^, with the exception of constipation, possibly due to imprecision inherent in record-based ascertainment. Most strikingly, the wearable-derived High-risk group achieved an LR+ of 7.91, approximately three times higher than that reported for questionnaire-based RBD screening in prior studies (LR+ = 2.8) ^13^, which underscores the utility of objective accelerometry over self-report measures. The apparent gap between the wearable-derived LR+ of 7.91 and the polysomnography-confirmed benchmark of 130 should be interpreted in context. The latter was derived from clinical and research cohorts of participants who may be in more symptomatic, thus advanced, disease stages, compared to community cohorts. In contrast, our classifier operated on an unselected population independent of RBD symptom status, representing a fundamentally earlier and broader point of intervention. Furthermore, the LR of 130 is anchored primarily to PD outcomes. When accounting for the full spectrum of α-synucleinopathy — including DLB in approximately 40% of iRBD cases, MSA in 5–10%, and a further proportion of participants potentially never reaching overt phenoconversion within any realistic follow-up window — the true predictive scope of iRBD, and by extension our wearable-derived signal, is likely underestimated. However, the UK Biobank’s linked health records do not allow reliable ascertainment of DLB or MSA outcomes for evaluation of the full synucleinopathy spectrum.

A key question was whether the wearable-derived RBD signal simply reflects inherited genetic susceptibility or captures a distinct, dynamic dimension of prodromal risk. Adjusting for PRS had minimal impact on the RBD association, with High-risk participants retaining approximately 4-fold elevated PD hazard after PRS adjustment (HR 4.42, 95% CI 2.86–6.84), suggesting that the wearable-derived signal may reflect environmental influences or rare genetic variants not captured by common variant PRS. Notably, PRS and RBD-classifier score each independently predicted PD, and their combination was synergistic. Participants with both elevated RBD-classifier scores and high genetic susceptibility faced amplified risk beyond either predictor alone (RBD × PD-PRS interaction HR 1.20 [95% CI 1.14–1.27]; p = 3.2 × 10^−10^; C-index 0.83). These findings are consistent with the two measures capturing complementary biological pathways, with one reflecting static inherited liability and the other ongoing neurophysiological change. *GBA1* analyses were underpowered (166 carriers; 4 incident PD cases), although a directionally elevated HR was observed (HR 2.10, 95% CI 0.99–4.44; p = 0.053).

While polysomnography-confirmed RBD remains among the strongest prodromal markers of PD [12], population-level screening with polysomnography is impractical. In contrast, wrist-worn accelerometry is inexpensive, passive, and scalable. The clinical utility of wearable-derived RBD scoring therefore lies not in standalone screening, but in risk enrichment. At the 90th percentile threshold, specificity was 91% while sensitivity was 34%, indicating that the score identifies a selectively enriched subgroup but does not capture most future PD cases. Complementary biomarkers will therefore be required to improve sensitivity while preserving specificity. The clinical utility of wearable-derived RBD scoring therefore lies not in standalone diagnosis, but in risk enrichment: identifying participants who may benefit from longitudinal monitoring, multimodal biomarker assessment, or enrollment into neuroprotective intervention trials. Importantly, the consistently high negative predictive value across thresholds is an expected consequence of the cohort’s low absolute PD incidence rather than a distinguishing property of the RBD score itself. In contrast, the positive predictive value increased approximately 6- to 15-fold over the cohort’s overall PD incidence across the 90th–99th percentile thresholds, supporting the use of the classifier for enrichment of a higher-risk subgroup. This framing also aligns with the regulatory environment for AI-derived wearable biomarkers, where risk communication and pre-specified actionability thresholds are required before clinical deployment ^31,32^.

At the most stringent threshold, 32 of 448 incident PD cases (7.1%) were identified within the top 1% of RBD-classifier scores, where 3.8% of participants developed PD. Thus, accelerometry provides substantial risk enrichment but limited sensitivity, supporting its use as a first-stage enrichment tool rather than a standalone screening strategy for prodromal neuroprotection trials. Further enrichment could be achieved by combining accelerometry symptoms-based RBD questionnaire and with complementary prodromal biomarkers, including olfactory testing and α-synuclein assays ^33,34^, before confirmatory DAT-SPECT ^35^. Such a staged approach could support more effective screening by reducing downstream burden and improving candidate selection, although prospective validation is required.

Several limitations should be considered. First, the wearable-derived RBD-classifier score cannot be validated against polysomnography in the UK Biobank and may capture sleep-related alterations beyond REM sleep without atonia, including wake and sleep fragmentation, periodic limb movements, and sleep apnea. Second, although selected prodromal and cognitive features were enriched in the High-risk group, individual prodromal markers provided limited independent predictive value for incident PD, additional to the test-retest reliability ^36^. This is consistent with their inherently modest LRs under MDS prodromal criteria — ranging from 1.6 for depression to 3.4 for erectile dysfunction ^13^ — and was likely further attenuated by derivation from hospital records, self-report, and medication use rather than structured clinical assessment. Third, cognitive measures utilized in the Cox regression analysis were not uniformly collected at the same visit as accelerometry; therefore, differences in assessment timing may have introduced measurement variability. Fourth, PD ascertainment relied on linked health records and may have missed undiagnosed or late-diagnosed cases. Fifth, the UK Biobank accelerometer cohort is not fully representative of the general population, and exclusions related to shift work or accelerometry quality may further limit generalizability. Sixth, despite strong relative risk estimates, absolute PD incidence remained low, limiting individual-level positive predictive value. Threshold-based sensitivity was also modest, limiting the use of the RBD-classifier score as a standalone screening tool, while exploratory *GBA1* analyses were underpowered. Finally, UK Biobank health record linkage does not support reliable ascertainment of DLB or MSA outcomes, limiting our ability to evaluate the full predictive scope of the wearable-derived signal across the α-synucleinopathy spectrum.

These findings establish wearable-derived nocturnal motor physiology as a scalable, passive, and biologically grounded signal for prodromal PD risk enrichment in population-based settings. Prior UK Biobank studies have shown that wrist accelerometry can characterize prodromal PD using activity-level, circadian rest-activity, or tremor-related features derived and modeled directly within the cohort itself. The present approach is conceptually distinct: it applies a classifier ^24^ that was developed and validated independently on clinical, polysomnography-confirmed RBD cases, without any retraining on PD outcomes or on this cohort, and demonstrates that this out-of-sample, disease-agnostic RBD signature nonetheless prospectively stratifies future PD risk at population scale. This RBD-centered approach complements previous passive, free-living wearable studies focused on tremor ^37^, 24-hour rest–activity rhythms ^38^ broader measures of mobility ^17^, and physical activity ^39,40^ in PD and its prodromal stages. Future work should integrate these complementary wrist-accelerometry modalities to define their respective and combined utility as population-level markers of prodromal PD risk. As neuroprotective trials increasingly depend on the enrollment of prodromal patients, population-scale wearable-based risk stratification may provide a scalable first-stage enrichment strategy toward identifying participants most likely to benefit from early intervention.

## METHODS

### Study Design and Population

We analyzed the UK Biobank prospective cohort, including participants who completed 7-day wrist accelerometry (Axivity AX3; 2013–2015) at baseline ^21,41^. After excluding prevalent neurological disease, poor-quality recordings, <4 valid recording nights, shift workers, prevalent Parkinson’s disease (PD), and incomplete linkage, 87,975 participants were included ^42,43^. Follow-up began at accelerometer deployment and continued until incident PD, death, or administrative censoring (30 November 2025). Non-converters were defined as participants without PD or dementia during follow-up.

The UK Biobank has approval from the North West Multicentre Research Ethics Committee as a Research Tissue Bank (RTB; reference: 21/NW/0157). This approval means that researchers do not require separate ethical clearance and can operate under the RTB approval. The UK Biobank obtained informed consent from all participants. This study was conducted under UK Biobank application number 97043 and in accordance with the Declaration of Helsinki.

### Outcome Ascertainment

Incident PD (fields 42030, 42031, 42032, 131022) was identified using UK Biobank algorithmically defined outcomes and linked Hospital Episode Statistics (HES) records (ICD-10 G20) ^44–46^. Alzheimer’s disease (42020, 42021, 131036), and vascular dementia (42022, 42023, 130838) were analyzed as disease-specific comparator outcomes using identical ascertainment procedures.

### Accelerometry-derived RBD-classifier score

Raw wrist accelerometry was processed using a previously developed and externally validated machine-learning classifier trained independently on polysomnography-confirmed idiopathic RBD (42 cases, 42 controls). Without retraining, the algorithm identified the principal sleep period, excluded invalid nights, extracted nocturnal movement features characteristic of REM sleep without atonia, and generated nightly RBD-classifier scores using gradient-boosted decision trees ^24,25^. Subject-level scores were obtained by averaging valid nights and stratified into Low (0–90th percentile), Intermediate (90–99th percentile), and High (99–100th percentile) risk groups.

Raw wrist accelerometry was processed using a previously developed and externally validated iRBD classifier ^19,24^. The classifier was developed in 42 polysomnography-confirmed iRBD patients and 42 controls, including 21 sleep-clinic patients with other sleep disorders and 21 community controls. After calibration, triaxial acceleration was converted to activity counts and summarized in 1-, 30-, and 60-second epochs. Automated sleep detection identified sleep periods, from which 119 features quantified activity index, twitch activity, short and long immobile bouts, mean motor activity, Hjorth parameters, and sleep timing and continuity across the sleep period and time windows enriched for REM sleep ^24^.

Features were entered into an ensemble of 400 boosted decision trees optimized with LogitBoost. Training and validation used subject-level leave-one-out cross-validation, with all nights from each held-out participant excluded during model fitting. The model generated a continuous nightly classification score, with positive values favoring iRBD and negative values favoring non-iRBD. Nightly scores were averaged across all valid nights to obtain a subject-level score. In the development cohort, the fully automated nighttime model achieved 95.2% sensitivity and 85.7% specificity at the subject level ^24^. Subsequent external validation across 352 iRBD and 258 non-RBD participants from four centers and multiple actigraphy devices yielded AUCs of 0.838–0.865 for the sleep model ^19^. In the UK Biobank, the pretrained model was applied without retraining; subject-level scores were averaged across valid nights and stratified into Low (0–90th percentile), Intermediate (90–99th percentile), and High (99–100th percentile) groups.

### Prodromal, Cognitive, and Genetic Markers

Prodromal markers included constipation (K59.0), depression (F32, F33, F34, F38, F39), anxiety (F40, F41), orthostatic hypotension (I95.1), erectile dysfunction (N52.01, N52.1, F52.21, N52.9), hyposmia (G52.0), and anosmia (R43.0), derived from UK Biobank questionnaires, medication records (Laxatives, Antidepressants, Anxiolytics, Vasopressors, PDE5 inhibitors), and HES data using adaptations of Movement Disorder Society prodromal criteria ^13,14^. Cognitive measures included fluid intelligence, reaction time, numeric memory, pairs matching, Trail Making Test (TMT)-A, TMT-B, and TMT-B/A ratio from assessments nearest the accelerometry visit ^47–53^. The TMT was introduced as part of the later enhanced cognitive assessment and was therefore unavailable for most participants at the original baseline visit, therefore an observational window of ±730 days of the accelerometry assessment were used.

Genetic analyses were performed in UK Biobank participants with available genotyping data from 487,410 participants included in the 2019 release. Genotypes were phased and imputed using the Haplotype Reference Consortium and UK10K/1000 Genomes reference panels ^54^. Sample-level quality control was performed using a GWAS pipeline as described previously ^55^. Polygenic risk scores (PRS) for PD and RBD were generated using the PRS-CS Bayesian continuous shrinkage framework based on published GWAS summary statistics and linkage disequilibrium reference panels ^56–58^.

*GBA1* variant status was determined from UK Biobank whole-genome sequencing data aligned to the GRCh38 reference genome using Gauchian, a locus-specific caller designed to accurately resolve variants, structural rearrangements, and recombinant alleles at the *GBA1* locus despite homology with the adjacent pseudogene *GBA1* ^59–61^.

### Statistical Analysis

Associations between accelerometry-derived RBD-classifier score and incident PD were estimated using Cox proportional hazards models 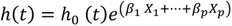 where *h(t)* denotes the hazard function at time *t*, *h*_0_(*t*) is the unspecified baseline hazard, and *β* represents regression coefficients. Tied event times were handled using the Breslow method (lifelines default) ^62–65^. Models were adjusted for age (21022), sex (31), BMI (23104), smoking status, and alcohol intake. Smoking (20117) and alcohol (20116) status was modeled as a three-level categorical variable (never/previous/current). RBD was modeled both continuously (per SD) and categorically (percentile groups). Hazard ratios (HRs) with 95% confidence intervals (CIs) and Harrell’s C-index were reported ^43,66–70^. Genetic analyses additionally adjusted for PD-PRS and the first ten ancestry principal components. Interaction models evaluated joint effects of RBD-classifier score with PD-PRS and GBA1 status ^43,66,67^.

Let *R* denote the RBD exposure, *P* the prodromal feature, and *X* the adjusted covariate vector. Their respective regression coefficients are denoted by *β*^R^, *β*^P^, *β*^X^, and *β*^RP^ for interaction effects. The baseline model 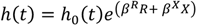 evaluated the total effect of RBD on incident disease. The model 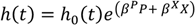 assessed the independent association of each prodromal feature. The additive model 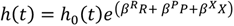 with the joint inclusion of RBD and prodromal features assessed their independent contributions. The multiplicative interaction model 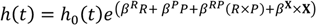 for effect modification was evaluated with cross-product interaction terms to evaluate relative risk due to interaction, attributable proportion and synergy index defined as RERI = *HR*_11_ − *HR*_10_ − *HR*_01_ + 1 with *HR*_10_ hazard ratios for RBD-only, *HR*_01_ prodromal-only, and *HR*_11_ the joint exposure. The Polygenic risk score adjustment model 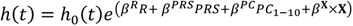 evaluated the association between PD, RBD-classifier score and PRS. The *GBA1* carrier interaction model 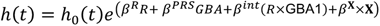 evaluated whether carrier status modified the association between RBD and incident disease. A competing risk model 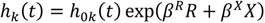 treated competing events as censoring. Competing events included incident Alzheimer’s disease, vascular dementia, and all-cause mortality occurring before PD diagnosis. Lastly, In the spline analysis, restricted cubic splines (RCS) with 4 degrees of freedom (3 internal knots) model the continuous dose–response relationship between RBD-classifier scores and log hazard. The reference point for the spline HR is the median RBD-classifier score. All Cox models employed sandwich variance estimators to obtain robust standard errors and included ridge penalization (λ = 0.01) for numerical stability^43,62,67,71–73^.

Sensitivity analyses evaluated competing-risk bias, 2-year lag exclusion from baseline to minimize reverse causality, RBD-classifier scores threshold robustness, and alternative model specifications ^43,63^. Competing-risk analyses incorporated cause-specific hazard modeling and Aalen–Johansen CIF vs. 1−KM. Additional analyses evaluated additive interaction metrics, absolute-risk stratification, and TMT subcohorts ^72^.

### Adjusted Comparison of Prodromal Features Across RBD score Strata on Non-Converters

Prodromal markers among non-converters were analyzed using multivariable logistic regression adjusted for age, sex, and follow-up duration. For each prodromal marker, a binary outcome was defined as presence at any assessment baseline or follow-up, where a value of 1 at either visit was coded as positive. Logistic regression was fitted separately for each marker using the form 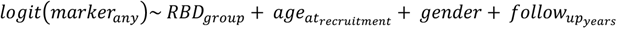.

Follow-up time was included as it represents differential observation window for symptom ascertainment. Odds ratios and 95% confidence intervals were derived from the exponentiated model coefficients. Models were fitted using maximum likelihood with up to 200 iterations. A global three-group chi-square test (unadjusted) was additionally computed per marker for comparison. Multiple testing correction used the Benjamini-Hochberg false discovery rate procedure applied across the five testable markers on the global chi-square p-values. All analyses were restricted to non-converters ^74,75^.

### Likelihood Ratio Screening

Stratum-specific likelihood ratios were computed for each RBD risk group^63^. For a given stratum *S* (e.g., High), 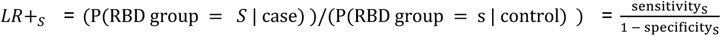 where sensitivity_S_ is the proportion of incident cases in stratum *S*, and specificity_S_ is the proportion of non-converters not in stratum *S*. The prior probability for Bayesian posterior computation was estimated empirically within 5-year age bands and interpolated by age, ranging from approximately 0.1% at age 50 to 2.2% at age 70. The cohort-wide 10-year PD incidence was 0.51% and is reported for context. Posterior probability was computed via the Fagan nomogram: post-test odds = pre-test odds × LR+, then converted to probability.

### Cross-Sectional Machine Learning Analysis

To evaluate the ability of accelerometry-derived RBD risk and established prodromal markers to identify participants who subsequently developed PD, we performed a cross-sectional machine learning analysis within the cohort. Participants with valid actigraphy recordings and complete covariate information were included, and models were trained to distinguish incident PD cases from non-converters using data available during the observational period prior to diagnosis. Predictor sets included demographics, accelerometry-derived RBD-classifier scores, prodromal features, PRS, and cognitive measures. Model performance was assessed using nested stratified cross-validation, with average receiver operating characteristic area under the curve (ROC-AUC) used to quantify discrimination. Models were trained cross-sectionally using baseline features but evaluated only on incident PD cases, making this an assessment of future PD identification rather than contemporaneous case classification.

### Software

Accelerometry-derived RBD-classifier scores were computed in MATLAB R2025b. All analyses were conducted in Python 3.13.9. Survival analyses, including Cox proportional hazards regression, Kaplan–Meier estimation, Aalen–Johansen cumulative incidence functions, Harrell’s C-index, and proportional hazards testing, were performed using lifelines 0.30.0 ^65,76^. Data manipulation and numerical operations were conducted using pandas 2.3.3 and NumPy 2.3.5, with SciPy 1.16.3 used for statistical testing, scikit-learn 1.7.2 for preprocessing, k-nearest-neighbor for bootstrap utilities, and statsmodels 0.14.5 for generalized linear models, Poisson regression sensitivity analyses, and false-discovery-rate correction. Intel MKL 2025.3.0 as the NumPy/SciPy linear algebra. The analysis pipeline used in this study is publicly available at ^77^

## Data Availability

The data that support the findings of this study are available from the UK Biobank under approved application, but restrictions apply to their availability because they were used under license for the current study and are therefore not publicly available. Researchers may obtain access directly from the UK Biobank by application.

## Code Availability

The analysis code used in this study is available in GitHub and can be accessed via this link https://github.com/GiorgioRicciardiello/UkbbRbdSleepPD-public.

## Acknowledgements

Not applicable.

## Author Contributions

G.R.M developed the statistical framework, implemented the computational pipeline, performed the survival, cognitive, and genetic interaction analyses, conducted literature review, interpreted the findings, and prepared the first manuscript draft. A.B-K. developed the accelerometry-derived RBD classifier and provided methodological guidance. L.L. contributed to the genetic analyses, interpretation of genetic data, and methodological guidance. L.Z., S.W. and A.P. contributed to data acquisition and manuscript revision. K.G. contributed to formal data analysis and manuscript revision. K.H.R. contributed to manuscript and figure preparation and scientific revisions. Z.G-O. provided expertise in Parkinson’s disease genetics, supervised the genetic analyses, interpreted the findings, and critically revised the manuscript. E.D. conceived the study, directed the project, supervised all stages of the work, interpreted the findings, provided scientific guidance throughout, and critically revised the manuscript. All authors read and approved the manuscript.

