## Supplementary Materials for "Accelerometry-Derived REM Sleep Behavior Disorder Score Is Associated with Incident Parkinson’s Disease in the UK Biobank"

#### Tables

| Description | Number Excluded | Percent | Number Retained |
|---|---|---|---|
| UKBB participants with valid actigraphy | | | 101,394 |
| No valid RBD-Classifier score (EHR — actigraphy merge) | 5876 | 5.8 | 95,500 |
| Pre-baseline neurological diagnosis | 1044 | 1.09 | 94,456 |
| Poor actigraphy quality | 5716 | 6.05 | 88,740 |
| Night-shift work at actigraphy time | 625 | 0.7 | 88,115 |
| Prevalent PD at actigraphy baseline | 140 | 0.20 | 87,975 |
| Final analytical cohort (Multi outcome) | | | 87,975 |

**Supplementary Table 1. Participant selection from the available dataset.** Accelerometry recordings were excluded based on UK Biobank quality-control variables indicating insufficient wear time (90015), failed or external calibration (90016–90017), daylight-savings crossover during recording (90018), unreliable device classification (90002), or recording-related technical issues (90180).

| Variable | Available N (%) | Overall (N=87,975) | Low (N=79,337) | Mid (N=7,800) | High (N=838) | p-value |
|---|---|---|---|---|---|---|
| **Demographics** | | | | | | |
| Age at recruitment (years) | 87,975 (100.0%) | 56.19 (7.80); 57.00 [50.00–62.00] | 55.87 (7.80); 57.00 [50.00–62.00] | 59.07 (7.19); 61.00 [55.00–65.00] | 59.60 (7.05); 61.00 [55.25–65.00] | <0.01 |
| Male sex | | 38367 (43.6%) | 32233 (40.6%) | 5505 (70.6%) | 629 (75.1%) | 0 |
| BMI (kg/m²) | | 26.70 (4.51); 26.02 [23.59–29.01] | 26.57 (4.45); 25.91 [23.49–28.86] | 27.75 (4.80); 27.02 [24.49–30.13] | 28.73 (5.37); 27.78 [25.16–31.24] | 1.43E-141 |
| **RBD-Classifier Score** | 87,975 (100.0%) | -1.85 (2.84); -2.18 [-3.95–-0.06] | -2.46 (2.23); -2.52 [-4.14–-0.78] | 3.46 (0.94); 3.29 [2.66–4.14] | 6.46 (0.66); 6.28 [5.93–6.86] | <0.01 |
| **Follow-up** | | | | | | |
| Follow-up time (years) | 87,975 (100.0%) | 9.98 (1.38); 10.20 [9.62–10.72] | 10.02 (1.32); 10.21 [9.64–10.72] | 9.69 (1.78); 10.04 [9.49–10.61] | 9.29 (2.19); 9.93 [9.34–10.45] | 4.897e-65 |
| **Incident Events (follow-up)** | | | | | | |
| Incident PD | - | 448 (0.5%) | 297 (0.4%) | 119 (1.5%) | 32 (3.8%) | <0.001 |
| Incident PD or AD | - | 29 (0.0%) | 21 (0.0%) | 5 (0.1%) | 3 (0.4%) | <0.001 |
| Incident Vascular Dementia | - | 122 (0.1%) | 96 (0.1%) | 21 (0.3%) | 5 (0.6%) | <0.001 |
| Incident PD + Vascular Dementia | - | 9 (0.0%) | 7 (0.0%) | 1 (0.0%) | 1 (0.1%) | 0.007 |
| Incident AD | - | 336 (0.4%) | 280 (0.4%) | 48 (0.6%) | 8 (1.0%) | <0.001 |
| **Cognitive Markers - Baseline** | | | | | | |
| Fluid Intelligence Score | 32,838 (37.3%) | 5.96 (1.97); 6.00 [5.00–7.00] | 5.97 (1.97); 6.00 [5.00–7.00] | 5.86 (2.03); 6.00 [4.00–7.00] | 5.85 (2.07); 6.00 [4.00–7.00] | 0.02124 |
| Reaction Time, ms | 87,736 (99.7%) | 545.04 (104.66); 527.00 [473.00–594.00] | 544.49 (104.55); 527.00 [473.00–593.00] | 549.59 (106.05); 531.00 [477.00–598.00] | 554.31 (100.32); 535.50 [485.00–598.00] | 8.521e-08 |
| Numeric Memory | 50,859 (57.8%) | 6.93 (1.47); 7.00 [6.00–8.00] | 6.94 (1.47); 7.00 [6.00–8.00] | 6.85 (1.50); 7.00 [6.00–8.00] | 6.71 (1.52); 7.00 [6.00–8.00] | 5.258e-06 |
| Pairs Matching Status | 53,994 (61.4%) | 0.03 (0.17); 0.00 [0.00–0.00] | 0.03 (0.16); 0.00 [0.00–0.00] | 0.04 (0.19); 0.00 [0.00–0.00] | 0.03 (0.17); 0.00 [0.00–0.00] | 4.969e-05 |
| TMT-A Duration, sec | 46,435 (52.8%) | 38.18 (13.53); 35.14 [28.88–44.07] | 38.01 (13.44); 35.01 [28.75–43.90] | 39.69 (14.30); 36.44 [30.11–45.59] | 41.00 (13.81); 36.77 [30.69–48.76] | 4.149e-18 |
| TMT-B Duration, sec | 46,435 (52.8%) | 66.59 (25.44); 61.04 [49.51–77.15] | 66.14 (25.24); 60.66 [49.20–76.53] | 70.74 (27.02); 64.31 [52.41–82.64] | 73.10 (26.05); 67.68 [54.17–87.06] | 1.748e-36 |
| TMT-B/A Ratio, log | 46,435 (52.8%) | 0.55 (0.26); 0.53 [0.37–0.71] | 0.55 (0.26); 0.53 [0.37–0.71] | 0.57 (0.26); 0.55 [0.39–0.73] | 0.57 (0.27); 0.55 [0.39–0.72] | 5.312e-07 |
| **Prodromal Markers - Baseline** | | | | | | |
| Constipation | 87,975 (100.0%) | 1962 (2.2%) | 1735 (2.2%) | 190 (2.4%) | 37 (4.4%) | 3.455e-05 |
| Depression | | 4629 (5.3%) | 3688 (4.6%) | 778 (10.0%) | 163 (19.5%) | 1.114e-162 |

| | | | | | | |
|---|---|---|---|---|---|---|
| Anxiety | | 383 (0.4%) | 320 (0.4%) | 54 (0.7%) | 9 (1.1%) | 1.998e-05 |
| Orthostatic Hypotension | | 153 (0.2%) | 126 (0.2%) | 24 (0.3%) | 3 (0.4%) | 0.004706 |
| Erectile Dysfunction | | 286 (0.3%) | 216 (0.3%) | 62 (0.8%) | 8 (1.0%) | 5.679e-16 |
| Dream Enactment | | 0 (0.0%) | 0 (0.0%) | 0 (0.0%) | 0 (0.0%) | — |
| Anosmia | | 5 (0.0%) | 5 (0.0%) | 0 (0.0%) | 0 (0.0%) | 0.7617 |
| Hyposmia | | 0 (0.0%) | 0 (0.0%) | 0 (0.0%) | 0 (0.0%) | — |
| **Prodromal Markers - Follow-up** | | | | | | |
| Constipation | | 2986 (3.4%) | 2584 (3.3%) | 352 (4.5%) | 50 (6.0%) | 7.497e-12 |
| Depression | | 1123 (1.3%) | 989 (1.2%) | 117 (1.5%) | 17 (2.0%) | 0.02451 |
| Anxiety | | 52 (0.1%) | 42 (0.1%) | 9 (0.1%) | 1 (0.1%) | 0.074 |
| Orthostatic Hypotension | 87,975 (100.0%) | 709 (0.8%) | 591 (0.7%) | 97 (1.2%) | 21 (2.5%) | 3.638e-12 |
| Erectile Dysfunction | | 149 (0.2%) | 125 (0.2%) | 23 (0.3%) | 1 (0.1%) | 0.01791 |
| Dream Enactment | | 0 (0.0%) | 0 (0.0%) | 0 (0.0%) | 0 (0.0%) | — |
| Anosmia | | 7 (0.0%) | 6 (0.0%) | 1 (0.0%) | 0 (0.0%) | 0.8547 |
| Hyposmia | | 0 (0.0%) | 0 (0.0%) | 0 (0.0%) | 0 (0.0%) | — |
| **Genetics** | | | | | | |
| PD PRS (z-score) | 66,952 (76.1%) | 0.02 (0.98); 0.03 [-0.64–0.69] | 0.02 (0.98); 0.02 [-0.64–0.68] | 0.04 (0.98); 0.06 [-0.61–0.71] | -0.01 (1.03); -0.01 [-0.74–0.67] | 0.0767 |
| RBD PRS (z-score) | 66,952 (76.1%) | 0.04 (0.90); 0.04 [-0.57–0.64] | 0.04 (0.90); 0.04 [-0.57–0.64] | 0.05 (0.91); 0.04 [-0.57–0.67] | -0.01 (0.89); 0.02 [-0.62–0.64] | 0.3876 |
| *GBA1* carrier | 70,693 (80.4%) | 167 (0.2%) | 152 (0.2%) | 14 (0.2%) | 1 (0.1%) | 0.8405 |

**Supplementary Table 2. Participant characteristics stratified by accelerometry-derived RBD risk group.** Characteristics of the analytical cohort (N = 87,975) stratified into Low (0–90th percentile), Intermediate (90–99th percentile), and High (99–100th percentile) accelerometry-derived RBD risk groups. Cognitive measures were obtained at assessments closest to the accelerometry recording and primarily reflect baseline cognitive performance. Prodromal features are presented separately for baseline and follow-up ascertainment to capture both prevalent and newly identified prodromal manifestations during longitudinal follow-up. Incident neurodegenerative outcomes were recorded over a median follow-up of approximately 10 years. Continuous variables are reported as mean (SD) and median [IQR], and categorical variables as n (%). P-values were calculated using Kruskal–Wallis tests for continuous variables and Pearson's $\chi^2$ or Fisher's exact tests for categorical variables, as appropriate. Increasing RBD risk was associated with greater prodromal burden, poorer cognitive performance, and higher incidence of Parkinson's disease.

| covariate | $\beta$ | SE($\beta$) | HR [95% CI] | z | p |
|---|---|---|---|---|---|
| $RBD_{High}$ | 1.546 | 0.194 | 4.69 [3.21, 6.87] | 7.95 | 1.80E-15 |
| $RBD_{Intermediate}$ | 0.628 | 0.087 | 1.87 [1.58, 2.22] | 7.18 | 6.81E-13 |
| Age | 0.034 | 0.004 | 1.03 [1.03, 1.04] | 9.31 | 1.29E-20 |
| Gender | 0.273 | 0.056 | 1.31 [1.18, 1.47] | 4.89 | 9.89E-07 |
| BMI | -0.002 | 0.006 | 1.00 [0.99, 1.01] | -0.38 | 7.05E-01 |
| Smoking | -0.021 | 0.044 | 0.98 [0.90, 1.07] | -0.46 | 6.45E-01 |
| Alcohol | -0.014 | 0.075 | 0.99 [0.85, 1.14] | -0.18 | 8.54E-01 |

**Supplementary Table 3. Baseline model RBD-only cox regression.** RBD as categorical variables adjusted by the age, gender, BMI, smoking, and alcohol. A total of 87,421 observations (controls and incident PD only) were included from which 448 were incident PD cases.

| Variable | Available N (%) | Overall (N=86,973) | Low (N=78,595) | Mid (N=7,593) | High (N=785) | p-value |
|---|---|---|---|---|---|---|
| **Cognitive Markers** | | | | | | |
| Fluid Intelligence Score | 32,495 (37.4%) | 5.96 (1.97); 6.00 [5.00–7.00] | 5.97 (1.97); 6.00 [5.00–7.00] | 5.87 (2.04); 6.00 [4.00–7.00] | 5.86 (2.08); 6.00 [4.00–7.00] | 0.03125 |
| Reaction Time, ms | 86,737 (99.7%) | 544.74 (104.49); 527.00 [473.00–594.00] | 544.20 (104.38); 527.00 [473.00–591.00] | 549.32 (106.00); 531.00 [477.00–598.00] | 554.18 (99.19); 536.00 [485.00–601.00] | 8.88e-08 |
| Numeric Memory | 50,359 (57.9%) | 6.94 (1.47); 7.00 [6.00–8.00] | 6.95 (1.46); 7.00 [6.00–8.00] | 6.87 (1.49); 7.00 [6.00–8.00] | 6.73 (1.52); 7.00 [6.00–8.00] | 2.665e-05 |
| Pairs Matching Status | 53,431 (61.4%) | 0.03 (0.17); 0.00 [0.00–0.00] | 0.03 (0.16); 0.00 [0.00–0.00] | 0.04 (0.19); 0.00 [0.00–0.00] | 0.03 (0.16); 0.00 [0.00–0.00] | 9.291e-05 |
| TMT-A Duration, sec | 45,988 (52.9%) | 38.11 (13.48); 35.08 [28.85–43.98] | 37.95 (13.39); 34.96 [28.73–43.84] | 39.54 (14.25); 36.30 [30.04–45.38] | 40.82 (13.56); 36.77 [30.58–48.84] | 4.269e-16 |
| TMT-B Duration, sec | 45,988 (52.9%) | 66.41 (25.27); 60.95 [49.44–76.97] | 65.99 (25.09); 60.57 [49.15–76.34] | 70.41 (26.73); 64.12 [52.22–82.26] | 72.89 (26.08); 67.55 [54.36–86.16] | 4.856e-34 |
| TMT-B/A Ratio, log | 45,988 (52.9%) | 0.55 (0.26); 0.53 [0.37–0.71] | 0.55 (0.26); 0.53 [0.37–0.70] | 0.57 (0.26); 0.55 [0.39–0.73] | 0.57 (0.27); 0.55 [0.39–0.72] | 3.656e-07 |
| **Prodromal Markers Baseline** | | | | | | |
| Constipation | 86,973 (100.0%) | 1924 (2.2%) | 1704 (2.2%) | 184 (2.4%) | 36 (4.6%) | 1.166e-05 |
| Depression | | 4567 (5.3%) | 3651 (4.6%) | 764 (10.1%) | 152 (19.4%) | 3.053e-158 |
| Anxiety | | 378 (0.4%) | 316 (0.4%) | 53 (0.7%) | 9 (1.1%) | 8.756e-06 |
| Orthostatic Hypotension | | 149 (0.2%) | 123 (0.2%) | 23 (0.3%) | 3 (0.4%) | 0.004659 |
| Erectile Dysfunction | | 276 (0.3%) | 211 (0.3%) | 60 (0.8%) | 5 (0.6%) | 3.209e-14 |
| Dream Enactment | | 0 (0.0%) | 0 (0.0%) | 0 (0.0%) | 0 (0.0%) | — |
| Anosmia | | 5 (0.0%) | 5 (0.0%) | 0 (0.0%) | 0 (0.0%) | 0.766 |
| Hyposmia | | 0 (0.0%) | 0 (0.0%) | 0 (0.0%) | 0 (0.0%) | — |
| **Prodromal - Follow-up** | | | | | | |
| Constipation | 86,973 (100.0%) | 2857 (3.3%) | 2484 (3.2%) | 329 (4.3%) | 44 (5.6%) | 3.802e-10 |
| Depression | | 1114 (1.3%) | 985 (1.3%) | 113 (1.5%) | 16 (2.0%) | 0.03658 |
| Anxiety | | 51 (0.1%) | 41 (0.1%) | 9 (0.1%) | 1 (0.1%) | 0.05387 |
| Orthostatic Hypotension | | 652 (0.7%) | 544 (0.7%) | 91 (1.2%) | 17 (2.2%) | 1.529e-10 |
| Erectile Dysfunction | | 149 (0.2%) | 125 (0.2%) | 23 (0.3%) | 1 (0.1%) | 0.01449 |
| Anosmia | | 6 (0.0%) | 5 (0.0%) | 1 (0.0%) | 0 (0.0%) | 0.7711 |

**Supplementary Table 4. Cognitive performance and prodromal features among non-converters stratified by accelerometry-derived RBD risk group.** Characteristics are shown for non-converters throughout follow-up (N = 86,973). Participants were stratified into Low (0–90th percentile), Intermediate (90–99th percentile), and High (99–100th percentile) accelerometry-derived RBD risk groups. Cognitive measures primarily reflect assessments obtained closest to the accelerometry visit, whereas prodromal features are presented separately for baseline and follow-up ascertainment. Continuous variables are reported as mean (SD) and median [IQR], and categorical variables as n (%). P-values were calculated using Kruskal–Wallis tests for continuous variables and Pearson's $\chi^2$ or Fisher's exact tests for categorical variables. Higher RBD risk was associated with progressively greater prodromal burden and poorer cognitive performance across multiple domains despite the absence of neurodegenerative disease diagnosis during follow-up. Abbreviation: TMT, Trail Making Test.

| | OR [95% CI] | | p-value | | | |
|---|---|---|---|---|---|---|
| **Marker** | **Mid** | **High** | **Mid** | **High** | **Global** | $p_{fdr}$ |
| Constipation | 1.14 [1.04-1.26] | 1.69 [1.33-2.15] | 0.0078 | 1.626e-05 | 6.389e-14 | 1.065e-13 |
| Depression | 2.88 [2.66-3.13] | 6.39 [5.34-7.63] | 9.876e-147 | 5.254e-92 | 1.118e-141 | 5.592e-141 |
| Anxiety | 1.91 [1.45-2.53] | 3.03 [1.60-5.73] | 5.478e-06 | 6.811e-04 | 6.968e-07 | 6.968e-07 |
| Orthostatic Hypotension | 1.19 [0.97-1.47] | 1.86 [1.18-2.94] | 0.0906 | 0.0077 | 9.794e-13 | 1.224e-12 |
| Erectile Dysfunction | 1.29 [1.01-1.65] | 0.82 [0.36-1.84] | 0.0413 | 0.6262 | 1.078e-14 | 2.696e-14 |

**Supplementary Table 5. Adjusted odds ratios for prodromal features at any assessment by accelerometry-derived RBD risk group among non-converters.** Values represent odds ratios (OR) with 95% confidence intervals (CI) from logistic regression models adjusted for age at recruitment, sex, and total follow-up time. The reference group is Low risk (0–90th percentile). Intermediate (90–99th percentile) and High (99–100th percentile) risk strata are compared against Low. N (%) columns reflect raw event count at any visit (baseline or follow-up) identical to Table 2. The global p-value is from an unadjusted three-group chi-square test. The FDR-adjusted p-value applies Benjamini-Hochberg correction across the five testable markers. Dream enactment, hyposmia, and anosmia were excluded due to structural zero event counts in Hospital Episode Statistics. Analyses were restricted to non-converters (N = 86,973).

| Outcome | covariate | $\beta$ SE($\beta$) | HR [95% CI] | z | p | N | events |
|---|---|---|---|---|---|---|---|
| Parkinson's Disease and Vascular Dementia | $RBD_{High}$ | 0.117 (0.357) | 1.12 [0.56, 2.26] | 0.329 | 0.742121 | 86805 | 9 |
| | $RBD_{Intermediate}$ | 0.003 (0.120) | 1.00 [0.79, 1.27] | 0.0264 | 0.978901 | | |
| | Age | 0.002 (0.004) | 1.00 [0.99, 1.01] | 0.4159 | 0.67747 | | |
| | Gender | 0.019 (0.068) | 1.02 [0.89, 1.16] | 0.2803 | 0.779281 | | |
| | BMI | 0.000 (0.007) | 1.00 [0.99, 1.01] | -0.0111 | 0.991144 | | |
| | Smoking | 0.002 (0.054) | 1.00 [0.90, 1.11] | 0.0308 | 0.975418 | | |
| | Alcohol | -0.002 (0.092) | 1.00 [0.83, 1.20] | -0.0232 | 0.981493 | | |
| Vascular Dementia | $RBD_{High}$ | 0.422 (0.325) | 1.52 [0.81, 2.88] | 1.2982 | 0.194219 | 86918 | 122 |
| | $RBD_{Intermediate}$ | 0.124 (0.112) | 1.13 [0.91, 1.41] | 1.1076 | 0.268038 | | |
| | Age | 0.017 (0.004) | 1.02 [1.01, 1.03] | 4.1525 | 3.29E-05 | | |
| | Gender | 0.079 (0.064) | 1.08 [0.95, 1.23] | 1.2363 | 0.216339 | | |
| | BMI | 0.006 (0.007) | 1.01 [0.99, 1.02] | 0.8055 | 0.420516 | | |
| | Smoking | 0.022 (0.051) | 1.02 [0.92, 1.13] | 0.4248 | 0.671017 | | |
| | Alcohol | -0.074 (0.086) | 0.93 [0.78, 1.10] | -0.8581 | 0.390814 | | |
| Parkinson's Disease and Alzheimer's Disease | $RBD_{High}$ | 0.340 (0.351) | 1.41 [0.71, 2.80] | 0.9705 | 0.331781 | 86825 | 29 |
| | $RBD_{Intermediate}$ | 0.035 (0.118) | 1.04 [0.82, 1.31] | 0.2956 | 0.767534 | | |
| | Age | 0.004 (0.004) | 1.00 [1.00, 1.01] | 0.9395 | 0.347454 | | |
| | Gender | 0.011 (0.067) | 1.01 [0.89, 1.15] | 0.1652 | 0.868795 | | |
| | BMI | -0.002 (0.007) | 1.00 [0.98, 1.01] | -0.28 | 0.779462 | | |
| | Smoking | -0.001 (0.054) | 1.00 [0.90, 1.11] | -0.0148 | 0.988154 | | |
| | Alcohol | 0.003 (0.091) | 1.00 [0.84, 1.20] | 0.0374 | 0.970127 | | |
| Alzheimer's Disease | $RBD_{High}$ | 0.412 (0.283) | 1.51 [0.87, 2.63] | 1.4573 | 0.145029 | 87130 | 334 |
| | $RBD_{Intermediate}$ | 0.155 (0.099) | 1.17 [0.96, 1.42] | 1.5561 | 0.119673 | | |
| | Age | 0.039 (0.004) | 1.04 [1.03, 1.05] | 10.3755 | 3.21E-25 | | |
| | Gender | 0.021 (0.058) | 1.02 [0.91, 1.14] | 0.359 | 0.719601 | | |
| | BMI | 0.001 (0.006) | 1.00 [0.99, 1.01] | 0.1801 | 0.857095 | | |
| | Smoking | 0.042 (0.046) | 1.04 [0.95, 1.14] | 0.9012 | 0.367509 | | |
| | Alcohol | -0.120 (0.076) | 0.89 [0.76, 1.03] | -1.5773 | 0.11473 | | |

**Supplementary Table 6. Model specificity for PD.** Cox regression model fitted for the different outcomes: Parkinson's and Vascular dementia, Vascular dementia, Parkinson's disease and Alzheimer's. Model were fitted with the adjusted covariates and the tertiles RBD risk groups. Only incident cases were considered as events.

| model | covariate | $\beta$ SE($\beta$) | HR [95% CI] | z | p | N | events |
| --- | --- | --- | --- | --- | --- | --- | --- |
| RBD Continuous with PRS | $RBD_z$ | 0.24 (0.03) | 1.27 [1.20, 1.35] | 7.8712 | 3.51E-15 | 66524 | 359 |
| | Age | 0.03 (0.00) | 1.04 [1.03, 1.04] | 8.2998 | 1.04E-16 | | |
| | Gender | 0.28 (0.06) | 1.32 [1.17, 1.50] | 4.3945 | 1.11E-05 | | |
| | BMI | 0.00 (0.01) | 1.00 [0.98, 1.01] | -0.3326 | 7.39E-01 | | |
| | Smoking | -0.02 (0.05) | 0.98 [0.88, 1.08] | -0.4803 | 6.31E-01 | | |
| | Alcohol | -0.06 (0.09) | 0.94 [0.79, 1.12] | -0.6824 | 4.95E-01 | | |
| | $PRS_{pd_z}$ | 0.31 (0.03) | 1.36 [1.28, 1.45] | 9.7184 | 2.52E-22 | | |
| | $pc1$ | 1.11 (5.22) | 3.04 [0.00, 84304.21] | 0.213 | 8.31E-01 | | |
| | $pc2$ | 0.13 (5.07) | 1.14 [0.00, 23394.71] | 0.0259 | 9.79E-01 | | |
| | $pc3$ | 1.18 (4.45) | 3.26 [0.00, 19842.22] | 0.2656 | 7.91E-01 | | |
| | $pc4$ | -1.59 (2.53) | 0.20 [0.00, 29.21] | -0.6285 | 5.30E-01 | | |
| | $pc5$ | 6.84 (5.19) | 929.88 [0.04, 24548074.92] | 1.3158 | 1.88E-01 | | |
| | $pc6$ | 1.66 (4.82) | 5.26 [0.00, 66283.07] | 0.3448 | 7.30E-01 | | |
| | $pc7$ | 4.43 (4.97) | 83.95 [0.01, 1416043.66] | 0.8921 | 3.72E-01 | | |
| | $pc8$ | 3.62 (3.16) | 37.17 [0.08, 18213.94] | 1.144 | 2.53E-01 | | |
| | $pc9$ | 2.21 (4.19) | 9.12 [0.00, 33578.48] | 0.5276 | 5.98E-01 | | |
| | $pc10$ | -0.07 (3.90) | 0.93 [0.00, 1953.35] | -0.0174 | 9.86E-01 | | |
| RBD Categorical with PRS | $RBD_{High}$ | 1.49 (0.22) | 4.42 [2.86, 6.84] | 6.676 | 2.46E-11 | 66524 | 359 |
| | $RBD_{Intermediate}$ | 0.61 (0.10) | 1.84 [1.52, 2.23] | 6.2091 | 5.33E-10 | | |
| | Age | 0.04 (0.00) | 1.04 [1.03, 1.04] | 8.4359 | 3.29E-17 | | |
| | Gender | 0.29 (0.06) | 1.33 [1.18, 1.51] | 4.5454 | 5.48E-06 | | |
| | BMI | 0.00 (0.01) | 1.00 [0.98, 1.01] | -0.2713 | 7.86E-01 | | |
| | Smoking | -0.02 (0.05) | 0.98 [0.88, 1.08] | -0.4605 | 6.45E-01 | | |
| | Alcohol | -0.06 (0.09) | 0.94 [0.79, 1.12] | -0.67 | 5.03E-01 | | |
| | $PRS_{pd_z}$ | 0.31 (0.03) | 1.36 [1.28, 1.45] | 9.717 | 2.55E-22 | | |
| | $pc1$ | 1.12 (5.22) | 3.06 [0.00, 85292.76] | 0.2145 | 8.30E-01 | | |
| | $pc2$ | 0.08 (5.06) | 1.08 [0.00, 22115.87] | 0.0155 | 9.88E-01 | | |
| | $pc3$ | 1.20 (4.45) | 3.31 [0.00, 20156.14] | 0.269 | 7.88E-01 | | |
| | $pc4$ | -1.60 (2.53) | 0.20 [0.00, 29.02] | -0.6312 | 5.28E-01 | | |
| | $pc5$ | 6.75 (5.19) | 854.88 [0.03, 22482931.64] | 1.3001 | 1.94E-01 | | |
| | $pc6$ | 1.82 (4.82) | 6.16 [0.00, 77888.03] | 0.3773 | 7.06E-01 | | |
| | $pc7$ | 4.46 (4.96) | 86.12 [0.01, 1448593.57] | 0.8975 | 3.69E-01 | | |
| | $pc8$ | 3.61 (3.16) | 37.02 [0.08, 18176.02] | 1.1423 | 2.53E-01 | | |
| | $pc9$ | 2.18 (4.19) | 8.82 [0.00, 32506.23] | 0.5197 | 6.03E-01 | | |
| | $pc10$ | 0.00 (3.90) | 1.00 [0.00, 2099.04] | 0.0009 | 9.99E-01 | | |

**Supplementary Table 7. Cox proportional hazards continuous RBD risk and PRS.** Cox Regression Hazard ratios (HRs) with 95% confidence intervals (CIs) and p-values are adjusted for age, sex, BMI, Smoking, alcohol use, PD-PRS, and the first 10 genetic principal components. Results demonstrate that the association between RBD-Classifier scores and incident PD remains robust after genetic adjustment, while PD-PRS independently predicts increased PD risk.

| covariate | $\beta$ SE($\beta$) | HR [95% CI] | z | p |
|---|---|---|---|---|
| $RBD_z$ | 0.21 (0.03) | 1.24 [1.17, 1.32] | 6.9039 | 5.06E-12 |
| $RBD_z \times PRS_{pd_z}$ | 0.18 (0.03) | 1.20 [1.14, 1.27] | 6.2905 | 3.17E-10 |
| Age | 0.03 (0.00) | 1.03 [1.03, 1.04] | 8.1241 | 4.51E-16 |
| Gender | 0.27 (0.06) | 1.30 [1.15, 1.48] | 4.1888 | 2.8E-05 |
| BMI | 0.00 (0.01) | 1.00 [0.98, 1.01] | -0.421 | 0.673732 |
| Smoking | -0.02 (0.05) | 0.98 [0.88, 1.08] | -0.466 | 0.641193 |
| Alcohol | -0.06 (0.09) | 0.94 [0.79, 1.12] | -0.6846 | 0.493586 |
| $PRS_{pd_z}$ | 0.28 (0.03) | 1.33 [1.25, 1.42] | 8.8258 | 1.09E-18 |
| $PRS_{rbd}$ | 0.03 (0.03) | 1.03 [0.96, 1.10] | 0.8565 | 0.391744 |
| $pc1$ | 1.11 (5.23) | 3.04 [0.00, 85217.64] | 0.2128 | 0.831488 |
| $pc2$ | 0.41 (5.08) | 1.51 [0.00, 31549.46] | 0.0811 | 0.935362 |
| $pc3$ | 1.05 (4.46) | 2.86 [0.00, 17805.85] | 0.2356 | 0.813753 |
| $pc4$ | -1.71 (2.54) | 0.18 [0.00, 26.62] | -0.6702 | 0.502715 |
| $pc5$ | 6.62 (5.20) | 752.40 [0.03, 20238840.62] | 1.2727 | 0.203123 |
| $pc6$ | 1.67 (4.82) | 5.33 [0.00, 67824.98] | 0.347 | 0.72857 |
| $pc7$ | 4.36 (4.98) | 78.10 [0.00, 1344946.31] | 0.8757 | 0.381188 |
| $pc8$ | 3.39 (3.17) | 29.58 [0.06, 14681.78] | 1.0696 | 0.28482 |
| $pc9$ | 1.96 (4.20) | 7.08 [0.00, 26471.47] | 0.4662 | 0.641091 |
| $pc10$ | 0.01 (3.91) | 1.01 [0.00, 2133.42] | 0.0016 | 0.998692 |

**Supplementary Table 8. Cox proportional hazards continuous RBD risk interaction with PRS**. Model evaluating the joint effect of continuous actigraphy-derived RBD-Classifier scores and Parkinson's disease (n=357) in the subset of 66,408 participants with available PRS. The model includes continuous RBD burden, PD-PRS, and their interaction term, adjusted for age, sex, BMI, Smoking, alcohol use, and the first 10 genetic principal components. Hazard ratios (HRs), 95% confidence intervals (CIs), and p-values are reported for the subset with available genetic data.

| covariate | $\beta$ SE($\beta$) | HR [95% CI] | z | p |
|---|---|---|---|---|
| $RBD_{High}$ | 1.21 (0.25) | 3.37 [2.06, 5.50] | 4.850 | 1.23E-06 |
| $RBD_{intermediate}$ | 0.50 (0.10) | 1.64 [1.35, 2.01] | 4.894 | 9.87E-07 |
| $PRS_{pd_z}$ | 0.26 (0.03) | 1.30 [1.22, 1.38] | 8.094 | 5.77E-16 |
| $RBD_{High} \times PRS_{pd_z}$ | 0.67 (0.17) | 1.96 [1.40, 2.74] | 3.911 | 9.19E-05 |
| $RBD_{intermediate} \times PRS_{pd_z}$ | 0.47 (0.09) | 1.60 [1.35, 1.90] | 5.376 | 7.62E-08 |
| Age | 0.03 (0.00) | 1.04 [1.03, 1.04] | 8.278 | 1.25E-16 |
| Gender | 0.28 (0.06) | 1.32 [1.16, 1.49] | 4.352 | 1.35E-05 |
| BMI | 0.00 (0.01) | 1.00 [0.98, 1.01] | -0.348 | 7.28E-01 |
| Smoking | -0.02 (0.05) | 0.98 [0.88, 1.08] | -0.460 | 6.45E-01 |
| Alcohol | -0.06 (0.09) | 0.94 [0.79, 1.13] | -0.662 | 5.08E-01 |
| $PRS_{pd_z}$ | 0.03 (0.03) | 1.03 [0.96, 1.10] | 0.812 | 4.17E-01 |
| $pc1$ | 1.28 (5.23) | 3.59 [0.00, 102117.57] | 0.244 | 8.07E-01 |
| $pc2$ | 0.39 (5.07) | 1.48 [0.00, 30859.35] | 0.077 | 9.38E-01 |
| $pc3$ | 1.08 (4.46) | 2.96 [0.00, 18497.75] | 0.243 | 8.08E-01 |
| $pc4$ | -1.79 (2.55) | 0.17 [0.00, 24.50] | -0.704 | 4.82E-01 |
| $pc5$ | 6.41 (5.20) | 607.37 [0.02, 16319061.40] | 1.232 | 2.18E-01 |
| $pc6$ | 1.87 (4.82) | 6.51 [0.00, 83227.15] | 0.388 | 6.98E-01 |
| $pc7$ | 4.44 (4.98) | 84.62 [0.00, 1455251.73] | 0.892 | 3.72E-01 |
| $pc8$ | 3.28 (3.17) | 26.50 [0.05, 13155.96] | 1.035 | 3.01E-01 |
| $pc9$ | 1.83 (4.20) | 6.25 [0.00, 23421.10] | 0.437 | 6.62E-01 |
| $pc10$ | 0.09 (3.91) | 1.10 [0.00, 2341.69] | 0.024 | 9.81E-01 |

**Supplementary Table 9. Cox proportional hazards categorical RBD risk interaction with PRS.** model evaluating whether PD polygenic risk modifies the association between categorical actigraphy-derived RBD strata and incident Parkinson's disease (n=357) in the subset of 66,408 participants with available PRS. Percentile-based RBD risk groups (Intermediate: 90–99th percentile; High: 99–100th percentile) were modeled together with continuous PD-PRS and corresponding interaction terms. Models were adjusted for demographic, lifestyle, and genetic ancestry covariates. Hazard ratios (HRs), 95% confidence intervals (CIs), and p-values are shown for the subset with available genetic data. Observations: 66,408; Events: 357.

**A. Continuous *GBA1* × RBD interaction Cox model**

| Variable | HR [95% CI] | p-value |
|---|---|---|
| RBD-Classifier score (per 1-SD) | 1.29 [1.22–1.37] | $1.1 \times 10^{-17}$ |
| *GBA1* carrier | 1.93 [0.65–5.73] | 0.236 |
| RBD × *GBA1* interaction | 2.10 [0.99–4.44] | 0.053 |
| Age (years) | 1.04 [1.03–1.04] | <0.001 |
| Gender (male) | 1.32 [1.17–1.49] | <0.001 |

Adjusted for BMI, Smoking, and alcohol use. N = 70,131; events = 388.
GBA1 carriers = 166; PD cases among carriers = 4. Harrell's C-index = 0.785; LRT interaction p = 0.076.

**B. Stratified event counts by RBD group and *GBA1* carrier status**

| RBD Group | $GBA^{-}$ N | $GBA^{-}$ PD events | $GBA^{+}$ N | $GBA^{+}$ PD events |
|---|---|---|---|---|
| $RBD_{low}$ | 63014 | 253 | 152 | 3 |
| $RBD_{intermediate}$ | 6275 | 103 | 13 | 0 |
| $RBD_{High}$ | 676 | 28 | 1 | 1 |

Sparse-event structure in *GBA1*+ strata resulted in quasi-complete separation in categorical interaction models.

**C. Categorical *GBA1* × RBD interaction model (exploratory)**

| Variable | HR 95% CI | p-value |
|---|---|---|
| $RBD_{intermediate}$ | 2.06 [1.72–2.48] | $9.9 \times 10^{-15}$ |
| $RBD_{High}$ | 0.82 [0.43–1.53] | 0.525 |
| $RBD_{intermediate}$× *GBA1* | 3.71 [0.02–852.0] | 0.636 |
| $RBD_{High}$× *GBA1* | Not estimable | - |

The $RBD_{High}$× *GBA1* interaction term could not be reliably estimated due to a single participant/event cell causing quasi-complete separation.

**Supplementary Table 10. Cox proportional hazards interaction with GBA1.** model evaluating whether GBA1 modifies the association between categorical actigraphy-derived RBD strata and incident Parkinson's disease (n=388). Hazard ratios (HRs), 95% confidence intervals (CIs), and p-values are shown for the subset with available genetic data. Observations: 66,408; Events: 357.

| Prodromal Variable | Covariate | $\beta$ SE($\beta$) | HR [95% CI] | z | p | N | events | $p_{fdr}$ |
|---|---|---|---|---|---|---|---|---|
| Fluid Intelligence | $Cognitive_{high}$ | -0.03 (0.12) | 0.97 [0.77, 1.22] | -0.23 | 8.11E-01 | 32668 | 173 | 0.8729 |
| | $Cognitive_{medium}$ | -0.02 (0.09) | 0.98 [0.82, 1.17] | -0.25 | 7.98E-01 | | | 0.8729 |
| | Age | 0.03 (0.01) | 1.03 [1.02, 1.05] | 5.78 | 7.28E-09 | | | |
| | Gender | 0.34 (0.09) | 1.41 [1.18, 1.68] | 3.77 | 1.59E-04 | | | |
| | BMI | 0.01 (0.01) | 1.01 [0.99, 1.03] | 0.55 | 5.79E-01 | | | |
| | Smoking | -0.03 (0.07) | 0.97 [0.84, 1.12] | -0.38 | 6.97E-01 | | | |
| | Alcohol | 0.01 (0.12) | 1.01 [0.80, 1.28] | 0.11 | 9.10E-01 | | | |
| Reaction Time (ms) | $Cognitive_{high}$ | 0.02 (0.06) | 1.02 [0.91, 1.14] | 0.28 | 7.72E-01 | 87185 | 448 | 0.8729 |
| | $Cognitive_{medium}$ | 0.04 (0.06) | 1.04 [0.92, 1.17] | 0.63 | 5.28E-01 | | | 0.8729 |
| | Age | 0.04 (0.00) | 1.04 [1.03, 1.04] | 9.72 | 2.29E-22 | | | |
| | Gender | 0.31 (0.06) | 1.37 [1.23, 1.52] | 5.642 | 1.68E-08 | | | |
| | BMI | 0.00 (0.01) | 1.00 [0.99, 1.01] | -0.04 | 9.64E-01 | | | |
| | Smoking | -0.01 (0.04) | 0.99 [0.91, 1.08] | -0.27 | 7.84E-01 | | | |
| | Alcohol | -0.02 (0.08) | 0.98 [0.85, 1.14] | -0.21 | 8.33E-01 | | | |
| Numeric Memory | $Cognitive_{high}$ | -0.17 (0.11) | 0.84 [0.67, 1.05] | -1.52 | 1.27E-01 | 50627 | 268 | 0.7400 |
| | $Cognitive_{medium}$ | -0.07 (0.07) | 0.94 [0.81, 1.08] | -0.92 | 3.57E-01 | | | 0.8729 |
| | Age | 0.04 (0.00) | 1.04 [1.03, 1.05] | 7.56 | 3.81E-14 | | | |
| | Gender | 0.30 (0.07) | 1.36 [1.18, 1.56] | 4.19 | 2.71E-05 | | | |
| | BMI | 0.00 (0.01) | 1.00 [0.98, 1.02] | -0.04 | 9.61E-01 | | | |
| | Smoking | -0.02 (0.06) | 0.98 [0.88, 1.10] | -0.29 | 7.69E-01 | | | |
| | Alcohol | -0.01 (0.10) | 0.99 [0.81, 1.22] | -0.05 | 9.55E-01 | | | |
| Pairs Matching Status | $Cognitive\ (yes)$ | -0.01 (0.21) | 0.99 [0.66, 1.48] | -0.05 | 9.56E-01 | 53717 | 286 | 0.9560 |
| | Age | 0.04 (0.00) | 1.04 [1.03, 1.05] | 7.96 | 1.67E-15 | | | |
| | Gender | 0.33 (0.07) | 1.38 [1.21, 1.59] | 4.62 | 3.77E-06 | | | |
| | BMI | 0.00 (0.01) | 1.00 [0.98, 1.02] | 0.01 | 9.90E-01 | | | |
| | Smoking | -0.02 (0.06) | 0.98 [0.88, 1.10] | -0.34 | 7.27E-01 | | | |
| | Alcohol | 0.03 (0.10) | 1.03 [0.85, 1.25] | 0.33 | 7.40E-01 | | | |
| Constipation | $Prodromal\ (yes)$ | 0.11 (0.18) | 1.12 [0.78, 1.60] | 0.59 | 5.52E-01 | 87421 | 448 | 0.8729 |
| | Age | 0.04 (0.00) | 1.04 [1.03, 1.04] | 9.76 | 1.61E-22 | | | |
| | Gender | 0.31 (0.06) | 1.37 [1.23, 1.52] | 5.63 | 1.73E-08 | | | |
| | BMI | 0.00 (0.01) | 1.00 [0.99, 1.01] | -0.04 | 9.61E-01 | | | |
| | Smoking | -0.01 (0.04) | 0.99 [0.91, 1.08] | -0.26 | 7.87E-01 | | | |
| | Alcohol | -0.01 (0.08) | 0.99 [0.85, 1.14] | -0.19 | 8.48E-01 | | | |
| Depression | $Prodromal\ (yes)$ | 0.07 (0.12) | 1.07 [0.84, 1.36] | 0.53 | 5.91E-01 | 87421 | 448 | 0.8729 |
| | Age | 0.04 (0.00) | 1.04 [1.03, 1.04] | 9.77 | 1.45E-22 | | | |
| | Gender | 0.31 (0.06) | 1.37 [1.23, 1.52] | 5.64 | 1.70E-08 | | | |
| | BMI | 0.00 (0.01) | 1.00 [0.99, 1.01] | -0.05 | 9.53E-01 | | | |
| | Smoking | -0.01 (0.04) | 0.99 [0.91, 1.08] | -0.27 | 7.84E-01 | | | |
| | Alcohol | -0.01 (0.08) | 0.99 [0.85, 1.14] | -0.18 | 8.49E-01 | | | |
| Anxiety | $Prodromal\ (yes)$ | -0.18 (0.43) | 0.84 [0.36, 1.93] | -0.42 | 6.74E-01 | 87421 | 448 | 0.8729 |

| | | | | | | | | |
|---|---|---|---|---|---|---|---|---|
| | Age | 0.04 (0.00) | 1.04 [1.03, 1.04] | 9.77 | 1.46E-22 | | | |
| | Gender | 0.31 (0.06) | 1.37 [1.23, 1.52] | 5.62 | 1.84E-08 | | | |
| | BMI | 0.00 (0.01) | 1.00 [0.99, 1.01] | -0.05 | 9.64E-01 | | | |
| | Smoking | -0.01 (0.04) | 0.99 [0.91, 1.08] | -0.26 | 7.90E-01 | | | |
| | Alcohol | -0.01 (0.08) | 0.99 [0.85, 1.14] | -0.19 | 8.43E-01 | | | |
| Orthostatic Hypotension | $Prodromal\ (yes)$ | 0.77 (0.55) | 2.16 [0.74, 6.32] | 1.40 | 1.59E-01 | 87421 | 448 | 0.7400 |
| | Age | 0.04 (0.00) | 1.04 [1.03, 1.04] | 9.75 | 1.68E-22 | | | |
| | Gender | 0.31 (0.06) | 1.37 [1.23, 1.52] | 5.62 | 1.89E-08 | | | |
| | BMI | 0.00 (0.01) | 1.00 [0.99, 1.01] | -0.04 | 9.64E-01 | | | |
| | Smoking | -0.01 (0.04) | 0.99 [0.91, 1.08] | -0.27 | 7.85E-01 | | | |
| | Alcohol | -0.01 (0.08) | 0.99 [0.85, 1.14] | -0.19 | 8.46E-01 | | | |
| Erectile Dysfunction | $Prodromal\ (yes)$ | 0.58 (0.40) | 1.79 [0.81, 3.95] | 1.43 | 1.50E-01 | 87421 | 448 | 0.7400 |
| | Age | 0.04 (0.00) | 1.04 [1.03, 1.04] | 9.74 | 1.88E-22 | | | |
| | Gender | 0.31 (0.06) | 1.36 [1.22, 1.52] | 5.58 | 2.32E-08 | | | |
| | BMI | 0.00 (0.01) | 1.00 [0.99, 1.01] | -0.05 | 9.53E-01 | | | |
| | Smoking | -0.01 (0.04) | 0.99 [0.90, 1.08] | -0.25 | 7.76E-01 | | | |
| | Alcohol | -0.01 (0.08) | 0.99 [0.85, 1.14] | -0.19 | 8.46E-01 | | | |

**Supplementary Table 11. Cox regression and prodromal association with incident PD.** Prodromal-only Cox proportional hazards models evaluating the association between individual prodromal features and incident Parkinson's disease. Each prodromal feature was modeled separately to estimate its independent association with future PD risk, adjusted for age, sex, BMI, Smoking, and alcohol use. Cognitive variables were analyzed using categorical groupings where applicable. Hazard ratios (HRs), 95% confidence intervals (CIs), raw p-values, and Benjamini–Hochberg false discovery rate (FDR)-adjusted p-values are reported. None of the tested prodromal features remained statistically significant after FDR correction.

| Variable | covariate | $\beta$ SE($\beta$) | HR [95% CI] | z | p | N | events |
|---|---|---|---|---|---|---|---|
| Fluid Intelligence | $RBD_{High}$ | 1.53 (0.33) | 4.63 [2.42, 8.84] | 4.64 | 3.50E-06 | 32668 | 173 |
| | $RBD_{Intermediate}$ | 0.75 (0.14) | 2.12 [1.60, 2.79] | 5.30 | 1.14E-07 | | |
| | $Cognitive_{high}$ | -0.03 (0.12) | 0.97 [0.77, 1.22] | -0.24 | 8.12E-01 | | |
| | $Prodromal_{medium}$ | -0.02 (0.09) | 0.98 [0.82, 1.17] | -0.24 | 8.12E-01 | | |
| | Age | 0.03 (0.01) | 1.03 [1.02, 1.04] | 5.46 | 4.79E-08 | | |
| | Gender | 0.30 (0.09) | 1.34 [1.13, 1.61] | 3.26 | 1.12E-03 | | |
| | BMI | 0.00 (0.01) | 1.00 [0.98, 1.02] | 0.33 | 7.42E-01 | | |
| | Smoking | -0.04 (0.07) | 0.96 [0.84, 1.11] | -0.52 | 6.04E-01 | | |
| | Alcohol | 0.02 (0.12) | 1.02 [0.81, 1.28] | 0.14 | 8.86E-01 | | |
| Reaction Time (ms) | $RBD_{High}$ | 1.55 (0.19) | 4.71 [3.22, 6.89] | 7.97 | 1.60E-15 | 87185 | 448 |
| | $RBD_{Intermediate}$ | 0.63 (0.09) | 1.87 [1.58, 2.23] | 7.19 | 6.70E-13 | | |
| | $Cognitive_{high}$ | 0.02 (0.06) | 1.02 [0.90, 1.14] | 0.26 | 7.96E-01 | | |
| | $Prodromal_{medium}$ | 0.04 (0.06) | 1.04 [0.92, 1.16] | 0.62 | 5.35E-01 | | |
| | Age | 0.03 (0.00) | 1.03 [1.03, 1.04] | 9.27 | 1.86E-20 | | |
| | Gender | 0.27 (0.06) | 1.31 [1.18, 1.47] | 4.90 | 9.37E-07 | | |
| | BMI | 0.00 (0.01) | 1.00 [0.99, 1.01] | -0.38 | 7.04E-01 | | |
| | Smoking | -0.02 (0.04) | 0.98 [0.90, 1.07] | -0.46 | 6.42E-01 | | |
| | Alcohol | -0.01 (0.08) | 0.99 [0.85, 1.14] | -0.20 | 8.44E-01 | | |
| Numeric Memory | $RBD_{High}$ | 1.81 (0.24) | 6.13 [3.80, 9.89] | 7.42 | 1.14E-13 | 50627 | 268 |
| | $RBD_{Intermediate}$ | 0.58 (0.12) | 1.79 [1.43, 2.26] | 4.99 | 6.10E-07 | | |
| | $Cognitive_{high}$ | -0.17 (0.11) | 0.84 [0.67, 1.06] | -1.47 | 1.42E-01 | | |
| | $Cognitive_{medium}$ | -0.06 (0.07) | 0.94 [0.81, 1.08] | -0.89 | 3.71E-01 | | |
| | Age | 0.03 (0.00) | 1.04 [1.03, 1.05] | 7.22 | 5.10E-13 | | |
| | Gender | 0.26 (0.07) | 1.30 [1.13, 1.50] | 3.61 | 3.04E-04 | | |
| | BMI | 0.00 (0.01) | 1.00 [0.98, 1.01] | -0.29 | 7.68E-01 | | |
| | Smoking | -0.02 (0.06) | 0.98 [0.87, 1.10] | -0.41 | 6.83E-01 | | |
| | Alcohol | 0.00 (0.10) | 1.00 [0.81, 1.22] | -0.05 | 9.62E-01 | | |
| Pairs Matching Status | $RBD_{High}$ | 1.78 (0.24) | 5.92 [3.71, 9.45] | 7.47 | 8.31E-14 | 53717 | 286 |
| | $RBD_{Intermediate}$ | 0.60 (0.11) | 1.83 [1.47, 2.28] | 5.36 | 8.22E-08 | | |
| | $Cognitive\ (yes)$ | -0.02 (0.21) | 0.98 [0.66, 1.47] | -0.08 | 9.39E-01 | | |
| | Age | 0.04 (0.00) | 1.04 [1.03, 1.05] | 7.60 | 2.85E-14 | | |
| | Gender | 0.28 (0.07) | 1.33 [1.16, 1.53] | 4.00 | 6.27E-05 | | |
| | BMI | 0.00 (0.01) | 1.00 [0.98, 1.01] | -0.26 | 7.94E-01 | | |
| | Smoking | -0.03 (0.06) | 0.97 [0.87, 1.09] | -0.48 | 6.31E-01 | | |
| | Alcohol | 0.03 (0.10) | 1.03 [0.85, 1.26] | 0.34 | 7.34E-01 | | |
| Constipation | $RBD_{High}$ | 1.54 (0.19) | 4.69 [3.20, 6.86] | 7.95 | 1.88E-15 | 87421 | 448 |
| | $RBD_{Intermediate}$ | 0.63 (0.09) | 1.87 [1.58, 2.22] | 7.18 | 6.82E-13 | | |
| | $Prodromal\ (yes)$ | 0.10 (0.18) | 1.10 [0.77, 1.58] | 0.54 | 5.89E-01 | | |
| | Age | 0.03 (0.00) | 1.03 [1.03, 1.04] | 9.30 | 1.37E-20 | | |
| | Gender | 0.27 (0.06) | 1.31 [1.18, 1.47] | 4.90 | 9.49E-07 | | |
| | BMI | 0.00 (0.01) | 1.00 [0.99, 1.01] | -0.38 | 7.03E-01 | | |

| | | | | | | | |
|---|---|---|---|---|---|---|---|
| | Smoking | -0.02 (0.04) | 0.98 [0.90, 1.07] | -0.46 | 6.44E-01 | | |
| | Alcohol | -0.01 (0.08) | 0.99 [0.85, 1.14] | -0.18 | 8.57E-01 | | |
| Depression | $RBD_{High}$ | 1.54 (0.19) | 4.69 [3.20, 6.86] | 7.94 | 2.09E-15 | 87421 | 448 |
| | $RBD_{Intermediate}$ | 0.63 (0.09) | 1.87 [1.58, 2.22] | 7.18 | 7.12E-13 | | |
| | $Prodromal\ (yes)$ | 0.01 (0.12) | 1.01 [0.80, 1.29] | 0.12 | 9.06E-01 | | |
| | Age | 0.03 (0.00) | 1.03 [1.03, 1.04] | 9.31 | 1.29E-20 | | |
| | Gender | 0.27 (0.06) | 1.31 [1.18, 1.47] | 4.90 | 9.82E-07 | | |
| | BMI | 0.00 (0.01) | 1.00 [0.99, 1.01] | -0.38 | 7.03E-01 | | |
| | Smoking | -0.02 (0.04) | 0.98 [0.90, 1.07] | -0.46 | 6.44E-01 | | |
| | Alcohol | -0.01 (0.08) | 0.99 [0.85, 1.14] | -0.18 | 8.55E-01 | | |
| Anxiety | $RBD_{High}$ | 1.55 (0.19) | 4.70 [3.21, 6.87] | 7.96 | 1.76E-15 | 87421 | 448 |
| | $RBD_{Intermediate}$ | 0.63 (0.09) | 1.87 [1.58, 2.22] | 7.19 | 6.70E-13 | | |
| | $Prodromal\ (yes)$ | -0.20 (0.43) | 0.82 [0.36, 1.88] | -0.48 | 6.35E-01 | | |
| | Age | 0.03 (0.00) | 1.03 [1.03, 1.04] | 9.31 | 1.26E-20 | | |
| | Gender | 0.27 (0.06) | 1.31 [1.18, 1.46] | 4.89 | 1.00E-06 | | |
| | BMI | 0.00 (0.01) | 1.00 [0.99, 1.01] | -0.38 | 7.05E-01 | | |
| | Smoking | -0.02 (0.04) | 0.98 [0.90, 1.07] | -0.46 | 6.47E-01 | | |
| | Alcohol | -0.01 (0.08) | 0.99 [0.85, 1.14] | -0.19 | 8.52E-01 | | |
| Orthostatic Hypotension | $RBD_{High}$ | 1.55 (0.19) | 4.70 [3.21, 6.87] | 7.96 | 1.76E-15 | 87421 | 448 |
| | $RBD_{Intermediate}$ | 0.63 (0.09) | 1.87 [1.58, 2.22] | 7.18 | 7.05E-13 | | |
| | $Prodromal\ (yes)$ | 0.76 (0.55) | 2.14 [0.73, 6.25] | 1.40 | 1.63E-01 | | |
| | Age | 0.03 (0.00) | 1.03 [1.03, 1.04] | 9.30 | 1.42E-20 | | |
| | Gender | 0.27 (0.06) | 1.31 [1.18, 1.46] | 4.89 | 1.02E-06 | | |
| | BMI | 0.00 (0.01) | 1.00 [0.99, 1.01] | -0.38 | 7.04E-01 | | |
| | Smoking | -0.02 (0.04) | 0.98 [0.90, 1.07] | -0.47 | 6.42E-01 | | |
| | Alcohol | -0.01 (0.08) | 0.99 [0.85, 1.14] | -0.18 | 8.56E-01 | | |
| Erectile Dysfunction | $RBD_{High}$ | 1.55 (0.19) | 4.69 [3.21, 6.87] | 7.95 | 1.80E-15 | 87421 | 448 |
| | $RBD_{Intermediate}$ | 0.63 (0.09) | 1.87 [1.58, 2.22] | 7.17 | 7.53E-13 | | |
| | $Prodromal\ (yes)$ | 0.55 (0.40) | 1.73 [0.79, 3.79] | 1.37 | 1.72E-01 | | |
| | Age | 0.03 (0.00) | 1.03 [1.03, 1.04] | 9.29 | 1.54E-20 | | |
| | Gender | 0.27 (0.06) | 1.31 [1.18, 1.46] | 4.86 | 1.20E-06 | | |
| | BMI | 0.00 (0.01) | 1.00 [0.99, 1.01] | -0.39 | 6.96E-01 | | |
| | Smoking | -0.02 (0.04) | 0.98 [0.90, 1.07] | -0.48 | 6.33E-01 | | |
| | Alcohol | -0.01 (0.08) | 0.99 [0.85, 1.14] | -0.18 | 8.56E-01 | | |

**Supplementary Table 12. Cox regression prodromal association and RDB risk with incident PD**. Additive Cox proportional hazards models jointly evaluating actigraphy-derived RBD risk strata and individual prodromal features for incident Parkinson's disease. Each prodromal feature was modeled separately together with categorical RBD risk groups (Low [reference], Intermediate [90th–99th percentile], High [99th–100th percentile]), adjusted for age, sex, BMI, Smoking, and alcohol use. Hazard ratios (HRs), 95% confidence intervals (CIs), and p-values are reported for both RBD strata and prodromal covariates. These models assess whether individual prodromal features contribute risk information beyond actigraphy-derived RBD burden.

| Prodromal Variable | covariate | $\beta$ SE($\beta$) | HR [95% CI] | z | p | N | events |
|---|---|---|---|---|---|---|---|
| Fluid Intelligence | $RBD_{high}$ | 1.18 (0.41) | 3.26 [1.46, 7.30] | 2.88 | 4.04E-03 | 32668 | 173 |
| | $RBD_{intermediate}$ | 0.65 (0.15) | 1.91 [1.42, 2.58] | 4.26 | 2.08E-05 | | |
| | $Cognitive_{high}$ | -0.04 (0.12) | 0.96 [0.76, 1.21] | -0.35 | 7.24E-01 | | |
| | $Cognitive_{medium}$ | -0.05 (0.09) | 0.95 [0.79, 1.14] | -0.55 | 5.82E-01 | | |
| | $RBD_{high} \times Cognitive_{high}$ | 0.57 (0.74) | 1.78 [0.42, 7.54] | 0.78 | 4.36E-01 | | |
| | $RBD_{high} \times Cognitive_{medium}$ | 0.88 (0.55) | 2.40 [0.81, 7.07] | 1.59 | 1.13E-01 | | |
| | $RBD_{intermediate} \times Cognitive_{high}$ | 0.31 (0.32) | 1.36 [0.73, 2.53] | 0.98 | 3.27E-01 | | |
| | $RBD_{intermediate} \times Cognitive_{medium}$ | 0.37 (0.22) | 1.44 [0.94, 2.22] | 1.67 | 9.56E-02 | | |
| | Age | 0.03 (0.01) | 1.03 [1.02, 1.04] | 5.42 | 6.08E-08 | | |
| | Gender | 0.29 (0.09) | 1.33 [1.11, 1.59] | 3.14 | 1.67E-03 | | |
| | BMI | 0.00 (0.01) | 1.00 [0.98, 1.02] | 0.30 | 7.61E-01 | | |
| | Smoking | -0.04 (0.07) | 0.96 [0.83, 1.11] | -0.54 | 5.87E-01 | | |
| | Alcohol | 0.02 (0.12) | 1.02 [0.81, 1.28] | 0.14 | 8.90E-01 | | |
| Reaction Time (ms) | $RBD_{high}$ | 1.40 (0.25) | 4.07 [2.51, 6.59] | 5.71 | 1.15E-08 | 87185 | 448 |
| | $RBD_{intermediate}$ | 0.51 (0.09) | 1.67 [1.39, 2.01] | 5.43 | 5.54E-08 | | |
| | $Cognitive_{high}$ | 0.01 (0.06) | 1.01 [0.90, 1.14] | 0.20 | 8.40E-01 | | |
| | $Cognitive_{medium}$ | 0.01 (0.06) | 1.01 [0.90, 1.14] | 0.20 | 8.42E-01 | | |
| | $RBD_{high} \times Cognitive_{high}$ | 0.19 (0.36) | 1.21 [0.60, 2.45] | 0.54 | 5.89E-01 | | |
| | $RBD_{high} \times Cognitive_{medium}$ | 0.42 (0.35) | 1.53 [0.77, 3.02] | 1.21 | 2.25E-01 | | |
| | $RBD_{intermediate} \times Cognitive_{high}$ | 0.24 (0.15) | 1.27 [0.96, 1.70] | 1.66 | 9.63E-02 | | |
| | $RBD_{intermediate} \times Cognitive_{medium}$ | 0.43 (0.14) | 1.54 [1.16, 2.04] | 2.97 | 2.94E-03 | | |
| | Age | 0.03 (0.00) | 1.03 [1.03, 1.04] | 9.13 | 7.13E-20 | | |
| | Gender | 0.27 (0.06) | 1.31 [1.17, 1.46] | 4.77 | 1.88E-06 | | |
| | BMI | 0.00 (0.01) | 1.00 [0.99, 1.01] | -0.44 | 6.57E-01 | | |
| | Smoking | -0.02 (0.04) | 0.98 [0.90, 1.07] | -0.51 | 6.09E-01 | | |
| | Alcohol | -0.01 (0.08) | 0.99 [0.85, 1.14] | -0.19 | 8.48E-01 | | |
| Numeric Memory | $RBD_{high}$ | 1.69 (0.31) | 5.41 [2.97, 9.87] | 5.50 | 3.70E-08 | 50627 | 268 |
| | $RBD_{intermediate}$ | 0.49 (0.13) | 1.64 [1.28, 2.09] | 3.92 | 8.79E-05 | | |
| | $Cognitive_{high}$ | -0.18 (0.12) | 0.84 [0.67, 1.05] | -1.51 | 1.31E-01 | | |
| | $Cognitive_{medium}$ | -0.09 (0.07) | 0.92 [0.79, 1.06] | -1.16 | 2.47E-01 | | |
| | $RBD_{high} \times Cognitive_{high}$ | 0.20 (0.92) | 1.22 [0.20, 7.38] | 0.21 | 8.32E-01 | | |
| | $RBD_{high} \times Cognitive_{medium}$ | 0.31 (0.39) | 1.36 [0.63, 2.94] | 0.78 | 4.33E-01 | | |
| | $RBD_{intermediate} \times Cognitive_{high}$ | 0.26 (0.34) | 1.30 [0.68, 2.51] | 0.79 | 4.30E-01 | | |
| | $RBD_{intermediate} \times Cognitive_{medium}$ | 0.32 (0.17) | 1.38 [1.00, 1.91] | 1.96 | 5.01E-02 | | |
| | Age | 0.03 (0.00) | 1.04 [1.03, 1.04] | 7.18 | 6.94E-13 | | |
| | Gender | 0.26 (0.07) | 1.29 [1.12, 1.49] | 3.50 | 4.59E-04 | | |
| | BMI | 0.00 (0.01) | 1.00 [0.98, 1.01] | -0.32 | 7.49E-01 | | |
| | Smoking | -0.03 (0.06) | 0.98 [0.87, 1.09] | -0.43 | 6.67E-01 | | |
| | Alcohol | 0.00 (0.10) | 1.00 [0.81, 1.22] | -0.05 | 9.61E-01 | | |
| | $RBD_{high}$ | 1.79 (0.24) | 6.01 [3.76, 9.59] | 7.50 | 6.40E-14 | 53717 | 286 |
| | $RBD_{intermediate}$ | 0.60 (0.11) | 1.82 [1.45, 2.27] | 5.28 | 1.30E-07 | | |

| | | | | | | | |
|---|---|---|---|---|---|---|---|
| Pairs Matching Status | $Cognitive_{high}$ | -0.03 (0.21) | 0.97 [0.64, 1.47] | -0.14 | 8.86E-01 | | |
| | $Cognitive_{medium}$ | -1.07 (2.01) | 0.34 [0.01, 17.63] | -0.53 | 5.94E-01 | | |
| | $RBD_{high} \times Cognitive_{high}$ | 0.28 (0.50) | 1.32 [0.49, 3.54] | 0.56 | 5.79E-01 | | |
| | $RBD_{high} \times Cognitive_{medium}$ | 0.04 (0.00) | 1.04 [1.03, 1.05] | 7.60 | 3.02E-14 | | |
| | $RBD_{intermediate} \times Cognitive_{high}$ | 0.28 (0.07) | 1.33 [1.16, 1.52] | 4.00 | 6.41E-05 | | |
| | $RBD_{intermediate} \times Cognitive_{medium}$ | 0.00 (0.01) | 1.00 [0.98, 1.01] | -0.25 | 8.02E-01 | | |
| | Smoking | -0.03 (0.06) | 0.97 [0.87, 1.09] | -0.48 | 6.30E-01 | | |
| | Alcohol | 0.03 (0.10) | 1.03 [0.85, 1.25] | 0.34 | 7.36E-01 | | |
| Constipation | $RBD_{high}$ | 1.56 (0.20) | 4.77 [3.25, 7.00] | 8.00 | 1.26E-15 | 87421 | 448 |
| | $RBD_{intermediate}$ | 0.63 (0.09) | 1.88 [1.58, 2.23] | 7.20 | 6.24E-13 | | |
| | $prodromal_{yes}$ | 0.13 (0.19) | 1.14 [0.79, 1.64] | 0.69 | 4.90E-01 | | |
| | $RBD_{high} \times prodromal_{yes}$ | -0.94 (1.20) | 0.39 [0.04, 4.14] | -0.78 | 4.35E-01 | | |
| | $RBD_{intermediate} \times prodromal_{yes}$ | -0.25 (0.56) | 0.78 [0.26, 2.32] | -0.45 | 6.54E-01 | | |
| | Age | 0.03 (0.00) | 1.03 [1.03, 1.04] | 9.30 | 1.37E-20 | | |
| | Gender | 0.27 (0.06) | 1.31 [1.18, 1.47] | 4.90 | 9.60E-07 | | |
| | BMI | 0.00 (0.01) | 1.00 [0.99, 1.01] | -0.38 | 7.05E-01 | | |
| | Smoking | -0.02 (0.04) | 0.98 [0.90, 1.07] | -0.46 | 6.47E-01 | | |
| | Alcohol | -0.01 (0.08) | 0.99 [0.85, 1.14] | -0.18 | 8.55E-01 | | |
| Depression | $RBD_{high}$ | 1.47 (0.21) | 4.36 [2.90, 6.53] | 7.12 | 1.08E-12 | 87421 | 448 |
| | $RBD_{intermediate}$ | 0.63 (0.09) | 1.87 [1.58, 2.23] | 7.12 | 1.11E-12 | | |
| | $prodromal_{yes}$ | -0.02 (0.13) | 0.98 [0.77, 1.26] | -0.13 | 8.96E-01 | | |
| | $RBD_{high} \times prodromal_{yes}$ | 0.51 (0.42) | 1.67 [0.73, 3.84] | 1.21 | 2.27E-01 | | |
| | $RBD_{intermediate} \times prodromal_{yes}$ | 0.05 (0.27) | 1.05 [0.61, 1.79] | 0.17 | 8.62E-01 | | |
| | Age | 0.03 (0.00) | 1.03 [1.03, 1.04] | 9.32 | 1.21E-20 | | |
| | Gender | 0.27 (0.06) | 1.31 [1.18, 1.47] | 4.90 | 9.53E-07 | | |
| | BMI | 0.00 (0.01) | 1.00 [0.99, 1.01] | -0.40 | 6.91E-01 | | |
| | Smoking | -0.02 (0.04) | 0.98 [0.90, 1.07] | -0.46 | 6.44E-01 | | |
| | Alcohol | -0.01 (0.08) | 0.99 [0.85, 1.14] | -0.17 | 8.66E-01 | | |
| Anxiety | $RBD_{high}$ | 1.55 (0.19) | 4.71 [3.22, 6.90] | 7.97 | 1.65E-15 | 87421 | 448 |
| | $RBD_{intermediate}$ | 0.63 (0.09) | 1.87 [1.58, 2.22] | 7.16 | 8.11E-13 | | |
| | $prodromal_{yes}$ | -0.24 (0.44) | 0.78 [0.33, 1.84] | -0.56 | 5.76E-01 | | |
| | $RBD_{high} \times prodromal_{yes}$ | -0.74 (2.54) | 0.48 [0.00, 69.30] | -0.29 | 7.72E-01 | | |
| | $RBD_{intermediate} \times prodromal_{yes}$ | 0.57 (0.93) | 1.76 [0.28, 10.97] | 0.61 | 5.45E-01 | | |
| | Age | 0.03 (0.00) | 1.03 [1.03, 1.04] | 9.31 | 1.27E-20 | | |
| | Gender | 0.27 (0.06) | 1.31 [1.18, 1.46] | 4.89 | 9.98E-07 | | |
| | BMI | 0.00 (0.01) | 1.00 [0.99, 1.01] | -0.38 | 7.05E-01 | | |
| | Smoking | -0.02 (0.04) | 0.98 [0.90, 1.07] | -0.46 | 6.46E-01 | | |
| | Alcohol | -0.01 (0.08) | 0.99 [0.85, 1.14] | -0.18 | 8.53E-01 | | |
| Orthostatic Hypotension | $RBD_{high}$ | 1.55 (0.19) | 4.70 [3.21, 6.88] | 7.96 | 1.73E-15 | 87421 | 448 |
| | $RBD_{intermediate}$ | 0.63 (0.09) | 1.88 [1.58, 2.23] | 7.19 | 6.30E-13 | | |
| | $prodromal_{yes}$ | 0.86 (0.57) | 2.35 [0.77, 7.16] | 1.51 | 1.32E-01 | | |
| | $RBD_{high} \times prodromal_{yes}$ | -0.86 (4.24) | 0.42 [0.0, 1739.5] | -0.20 | 8.40E-01 | | |
| | $RBD_{intermediate} \times prodromal_{yes}$ | -0.88 (1.54) | 0.41 [0.02, 8.49] | -0.57 | 5.66E-01 | | |

| | | | | | | | |
|---|---|---|---|---|---|---|---|
| | Age | 0.03 (0.00) | 1.03 [1.03, 1.04] | 9.30 | 1.42E-20 | | |
| | Gender | 0.27 (0.06) | 1.31 [1.18, 1.46] | 4.89 | 1.01E-06 | | |
| | BMI | 0.00 (0.01) | 1.00 [0.99, 1.01] | -0.38 | 7.05E-01 | | |
| | Smoking | -0.02 (0.04) | 0.98 [0.90, 1.07] | -0.46 | 6.43E-01 | | |
| | Alcohol | -0.01 (0.08) | 0.99 [0.85, 1.14] | -0.18 | 8.54E-01 | | |
| Erectile Dysfunction | $RBD_{high}$ | 1.52 (0.20) | 4.57 [3.11, 6.72] | 7.74 | 1.03E-14 | 87421 | 448 |
| | $RBD_{intermediate}$ | 0.63 (0.09) | 1.87 [1.58, 2.22] | 7.17 | 7.73E-13 | | |
| | $prodromal_{yes}$ | 0.42 (0.44) | 1.53 [0.64, 3.63] | 0.96 | 3.37E-01 | | |
| | $RBD_{high} \times prodromal_{yes}$ | 1.69 (1.12) | 5.43 [0.61, 48.42] | 1.52 | 1.30E-01 | | |
| | $RBD_{intermediate} \times prodromal_{yes}$ | 0.02 (0.84) | 1.02 [0.20, 5.26] | 0.02 | 9.83E-01 | | |
| | age | 0.03 (0.00) | 1.03 [1.03, 1.04] | 9.30 | 1.44E-20 | | |
| | Gender | 0.27 (0.06) | 1.31 [1.18, 1.46] | 4.85 | 1.21E-06 | | |
| | BMI | 0.00 (0.01) | 1.00 [0.99, 1.01] | -0.40 | 6.90E-01 | | |
| | Smoking | -0.02 (0.04) | 0.98 [0.90, 1.07] | -0.48 | 6.30E-01 | | |
| | Alcohol | -0.01 (0.08) | 0.99 [0.85, 1.14] | -0.17 | 8.62E-01 | | |

**Supplementary Table 13. Cox regression prodromal association and RDB risk interaction with incident PD.** Interaction Cox proportional hazards models evaluating whether individual prodromal features modify the association between actigraphy-derived RBD burden and incident Parkinson's disease. Each model included categorical RBD risk strata (Low [reference], Intermediate [90th–99th percentile], High [99th–100th percentile]), an individual prodromal feature, and corresponding interaction terms between RBD strata and prodromal categories. Models were adjusted for age, sex, BMI, Smoking, and alcohol use. Hazard ratios (HRs), 95% confidence intervals (CIs), and p-values are reported for main and interaction effects. These analyses assess whether prodromal features alter the predictive relationship between RBD burden and future PD incidence.

| Prodromal feature | IRR (RBD only) | IRR (Prodromal only) | IRR (Joint Exposure) | RERI | Synergy Index | Jointly Exposed N | Jointly Exposed Events |
|---|---|---|---|---|---|---|---|
| Constipation | 5.5 | 1.37 | 0 | -5.86 | -0.21 | 36 | 0 |
| Depression | 5.02 | 1.25 | 7.3 | 2.04 | 1.48 | 158 | 6 |
| Anxiety | 5.33 | 0.52 | 0 | -4.85 | -0.26 | 9 | 0 |
| Orthostatic Hypotension | 5.32 | 2.84 | 0 | -7.16 | -0.16 | 3 | 0 |
| Erectile Dysfunction | 5.18 | 1.49 | 31.74 | 26.08 | 6.59 | 6 | 1 |

**Supplementary Table 14. Additive interaction with RBD risk for incident Parkinson's.** Additive interaction analyses between actigraphy-derived RBD burden and prodromal features (binary, yes/no events) for incident Parkinson's disease using Poisson regression sensitivity models. Incidence rate ratios (IRRs) are reported for isolated RBD exposure ($IRR_{10}$), isolated prodromal feature exposure ($IRR_{01}$), and joint exposure ($IRR_{11}$). Additive interaction was quantified using the Relative Excess Risk due to Interaction (RERI), attributable proportion (AP), and synergy index (SI). Poisson regression models included log(time) offsets and were used as sensitivity analyses to improve estimate stability under sparse-event conditions, with IRRs approximating hazard ratios under the rare-disease assumption. Jointly exposed subgroup counts and event frequencies are provided to facilitate interpretation of sparse or zero-event strata, which may yield unstable interaction estimates.

**A. Cognitive performance**

| RBD Group | Cognitive Test | Cognitive Category | 10-Year PD Incidence % (95% CI) | N | Events |
|---|---|---|---|---|---|
| High | Numeric Memory | Low | 7.03 (3.95–12.33) | 163 | 11 |
| High | Reaction Time | Low | 6.73 (3.97–11.27) | 234 | 14 |
| High | Pairs Matching | Normal | 5.26 (3.50–7.89) | 441 | 22 |
| High | Fluid Intelligence Score | Medium | 5.02 (2.12–11.64) | 105 | 5 |
| High | Fluid Intelligence Score | Low | 4.93 (2.50–9.63) | 173 | 8 |
| Intermediate | Reaction Time | Medium | 1.89 (1.40–2.53) | 2547 | 44 |
| Low | Numeric Memory | Low | 0.53 (0.42–0.65) | 15847 | 81 |

**B. Prodromal features**

| RBD Group | Prodromal feature | Prodromal Category | 10-Year PD Incidence % (95% CI) | N | Events |
|---|---|---|---|---|---|
| High | Erectile Dysfunction | Yes | 16.67 (2.53–72.69) | 6 | 1 |
| High | Constipation | No | 4.43 (3.14–6.23) | 781 | 32 |
| High | Depression | No | 4.34 (2.96–6.35) | 659 | 26 |
| High | Anxiety | No | 4.28 (3.03–6.03) | 808 | 32 |
| High | Orthostatic Hypotension | No | 4.25 (3.01–5.98) | 814 | 32 |
| High | Erectile Dysfunction | No | 4.14 (2.92–5.87) | 811 | 31 |
| Intermediate | Depression | Yes | 0.96 (0.46–2.00) | 771 | 7 |
| Low | Orthostatic Hypotension | Yes | 2.61 (0.85–7.88) | 126 | 3 |

**Supplementary Table 15. Absolute cumulative incidence of Parkinson's at Ten-year Kaplan–Meier cumulative incidence.** PD across combined actigraphy-derived RBD and prodromal risk strata. Ten-year Kaplan–Meier cumulative incidence estimates with 95% confidence intervals are reported for joint RBD–prodromal subgroups, including cognitive performance and prodromal features. Cognitive variables were categorized into tertiles (Low, Medium, High) using quantile-based binning, while binary prodromal variables were categorized as present or absent. Estimates derived from very small jointly exposed groups should be interpreted cautiously.

**A. Prodromal-only TMT model (Model 1)**

| Variable | HR [95% CI] | p-value |
|---|---|---|
| $cognitive_{high}$ | 0.95 [0.81, 1.11] | 5.20E-01 |
| $cognitive_{medium}$ | 0.97 [0.83, 1.14] | 7.09E-01 |
| Age | 1.04 [1.03, 1.05] | 2.98E-12 |
| Gender | 1.35 [1.16, 1.57] | 8.53E-05 |
| BMI | 1.00 [0.98, 1.02] | 9.81E-01 |
| Smoking | 0.98 [0.87, 1.11] | 7.42E-01 |
| Alcohol | 0.98 [0.80, 1.21] | 8.52E-01 |

**B. Additive TMT + RBD model (Model 2)**

| Variable | HR [95% CI] | p-value |
|---|---|---|
| $RBD_{high}$ | 5.25 [3.05, 9.04] | 2.10E-09 |
| $RBD_{intermediate}$ | 1.85 [1.45, 2.35] | 5.21E-07 |
| $cognitive_{high}$ | 0.95 [0.81, 1.11] | 4.99E-01 |
| $cognitive_{medium}$ | 0.97 [0.83, 1.14] | 7.05E-01 |
| Age | 1.03 [1.02, 1.04] | 2.44E-11 |
| Gender | 1.30 [1.12, 1.51] | 6.67E-04 |
| BMI | 1.00 [0.98, 1.01] | 8.01E-01 |
| Smoking | 0.97 [0.86, 1.10] | 6.66E-01 |
| Alcohol | 0.98 [0.80, 1.21] | 8.57E-01 |

**C. Interaction TMT × RBD model (Model 3)**

| Variable | HR [95% CI] | p-value |
|---|---|---|
| $RBD_{high}$ | 3.59 [1.77, 7.28] | 3.95E-04 |
| $RBD_{intermediate}$ | 1.71 [1.32, 2.22] | 4.70E-05 |
| $cognitive_{high}$ | 0.95 [0.81, 1.12] | 5.33E-01 |
| $cognitive_{medium}$ | 0.94 [0.80, 1.11] | 4.55E-01 |
| $RBD_{high} \times Cognitive_{high}$ | 1.81 [0.67, 4.91] | 2.41E-01 |
| $RBD_{high} \times Cognitive_{medium}$ | 2.30 [0.87, 6.08] | 9.40E-02 |
| $RBD_{intermediate} \times Cognitive_{high}$ | 1.05 [0.70, 1.57] | 8.05E-01 |
| $RBD_{intermediate} \times Cognitive_{medium}$ | 1.52 [1.02, 2.26] | 4.07E-02 |
| Age | 1.03 [1.02, 1.04] | 3.78E-11 |
| Gender | 1.29 [1.11, 1.50] | 9.91E-04 |
| BMI | 1.00 [0.98, 1.01] | 7.62E-01 |
| Smoking | 0.97 [0.86, 1.10] | 6.49E-01 |
| Alcohol | 0.98 [0.79, 1.21] | 8.44E-01 |

**Supplementary Table 16. Trail Making Test B/A (TMT-B/A) ratio as a measure of executive function.** Analyses were restricted to the subcohort with available online cognitive assessment data within ±730 days of baseline actigraphy (N = 46,227; 239 incident PD cases). Model 1 evaluated the independent association between TMT performance and incident Parkinson’s disease. Model 2 jointly evaluated TMT categories and actigraphy-derived RBD burden, while Model 3 assessed multiplicative interaction effects between TMT performance and RBD strata. All Cox proportional hazards models were adjusted for age, sex, BMI, Smoking, and alcohol use.

**A. CIF vs KM**

| RBD Group | Time Horizon (Years) | Aalen–Johansen CIF (%) | Kaplan–Meier Estimate (%) |
|---|---|---|---|
| High (99,100%) | 5 | 2.26 | 2.26 |
| | 10 | 4.24 | 4.24 |
| Intermediate (90,99%) | 5 | 0.62 | 0.62 |
| | 10 | 1.65 | 1.65 |
| Low (0,90%) | 5 | 0.16 | 0.16 |
| | 10 | 0.38 | 0.38 |

**B. Cause Specific Cox**

| Covariate | HR [95% CI] | p-value |
|---|---|---|
| $RBD_{High}$ | 4.69 [3.21, 6.87] | 1.80E-15 |
| $RBD_{intermediate}$ | 1.87 [1.58, 2.22] | 6.81E-13 |
| Age | 1.03 [1.03, 1.04] | 1.29E-20 |
| Gender | 1.31 [1.18, 1.47] | 9.89E-07 |
| BMI | 1.00 [0.99, 1.01] | 7.05E-01 |
| Smoking | 0.98 [0.90, 1.07] | 6.45E-01 |
| Alcohol | 0.99 [0.85, 1.14] | 8.54E-01 |

**Supplementary Table 17. Competing-risk analyses for incident Parkinson's disease. (A)** Aalen–Johansen cumulative incidence function (CIF) estimates accounting for competing events (Alzheimer's disease, vascular dementia, and all-cause death) compared with standard Kaplan–Meier estimates across actigraphy-derived RBD strata and follow-up horizons. Absolute differences quantify the magnitude of competing-risk bias. **(B)** Cause-specific Cox proportional hazards models evaluating the association between actigraphy-derived RBD burden and incident Parkinson's disease while treating competing neurological diagnoses and death as censored events. Models were adjusted for age, sex, BMI, Smoking, and alcohol consumption. Hazard ratios (HRs), 95% confidence intervals (CIs), p-values, concordance statistics (C-index), sample sizes, and incident PD events are reported.

| Percentile | Threshold Value | HR [95% CI] | Number high | Number low | p |
|---|---|---|---|---|---|
| 5 | 3.4 | 2.51 [2.05, 3.01] | 4372 | 83049 | 9.08E-19 |
| 10 | 2.1 | 2.01 [1.77, 2.44] | 8743 | 78678 | 2.28E-19 |
| 15 | 1.2 | 1.80 [1.56, 2.01] | 13114 | 74307 | 2.23E-16 |

**Supplementary Table 18. RBD Threshold sensitivity**. Cox regression was tested at different RBD risk group threshold for sensitivity analysis. Hazards ratio for the high RBD risk group is presented alongside the number of participants in the respective high and low strata.

| Percentile | Time Horizon (Years) | RBD | Number Evaluated | PD Incident Rate | Sensitivity [95% CI] | Specificity [95% CI] | PPV [95% CI] | NPV [95% CI] | TP | FP | FN | TN |
|---|---|---|---|---|---|---|---|---|---|---|---|---|
| 90 | 5 | 2.1 | 85678 | 0.002253 | 0.34 [0.28-0.41] | 0.90 [0.89, 0.91] | 0.01 [0.0061-0.0098] | 1.00 [0.9980-0.9986] | 65 | 8355 | 128 | 77130 |
| | 10 | | 50816 | 0.008737 | 0.34 [0.30-0.39] | 0.91 [0.90-0.91] | 0.03 [0.0279-0.0381] | 0.99 [0.9929-0.9944] | 152 | 4511 | 292 | 45861 |
| 95 | 5 | 3.4 | 85678 | 0.002253 | 0.21 [0.17-0.28] | 0.95 [0.95-0.96] | 0.01 [0.0072-0.0133] | 1.00 [0.9978-0.9984] | 41 | 4145 | 152 | 81340 |
| | 10 | | 50816 | 0.008737 | 0.22 [0.190.26] | 0.96 [0.95-0.96] | 0.04 [0.0355-0.0523] | 0.99 [0.9921-0.9936] | 98 | 2175 | 346 | 48197 |
| 99 | 5 | 5.6 | 85678 | 0.002253 | 0.09 [0.06-0.15] | 0.99 [0.98-0.99] | 0.02 [0.0138-0.0340] | 1.00 [0.9976-0.9982] | 18 | 812 | 175 | 84673 |
| | 10 | | 50816 | 0.008737 | 0.07 [0.05-0.11] | 0.99 [0.99-0.99] | 0.07 [0.0537-0.1030] | 0.99 [0.9910-0.9926] | 33 | 409 | 411 | 49963 |

**Supplementary Table 19. Predictive performance of actigraphy-derived RBD probability thresholds for incident Parkinson's disease at 5- and 10-year follow-up horizons.** Percentile-based thresholds (90th, 95th, and 99th percentiles) were evaluated as binary screening cutoffs. Reported metrics include sensitivity, specificity, positive predictive value (PPV), negative predictive value (NPV), and confusion matrix counts (TP, FP, FN, TN) with 95% confidence intervals. Increasing threshold stringency improved specificity and PPV at the cost of reduced sensitivity, while NPV remained >99% across all thresholds.

| Marker | Category | HR (Primary Model) | HR (2-Year Lag Model) | P - value | N | Events |
|---|---|---|---|---|---|---|
| Fluid Intelligence | $cognitive_{high}$ | 0.97 | 0.99 [0.78, 1.24] | 9.09E-01 | 32651 | 156 |
| | $cognitive_{medium}$ | 0.97 | 0.98 [0.81, 1.17] | 7.95E-01 | | |
| Reaction Time (ms) | $cognitive_{high}$ | 1.02 | 1.03 [0.91, 1.16] | 6.24E-01 | 87134 | 397 |
| | $cognitive_{medium}$ | 1.02 | 1.04 [0.92, 1.17] | 5.31E-01 | | |
| Numeric Memory | $cognitive_{high}$ | 0.84 | 0.83 [0.66, 1.04] | 1.03E-01 | 50599 | 240 |
| | $cognitive_{medium}$ | 0.84 | 0.94 [0.82, 1.09] | 4.21E-01 | | |
| Pairs Matching Status | $prodromal_{yes}$ | 0.99 | 0.99 [0.66, 1.49] | 9.61E-01 | 53687 | 256 |
| Constipation | $prodromal_{yes}$ | 1.12 | 1.13 [0.78, 1.63] | 5.24E-01 | | |
| Depression | $prodromal_{yes}$ | 1.07 | 1.01 [0.79, 1.30] | 9.29E-01 | | |
| Anxiety | $prodromal_{yes}$ | 0.84 | 0.87 [0.37, 2.03] | 7.42E-01 | 87370 | 397 |
| Orthostatic Hypotension | $prodromal_{yes}$ | 2.16 | 2.33 [0.78, 6.95] | 1.28E-01 | | |
| Erectile Dysfunction | $prodromal_{yes}$ | 1.79 | 1.93 [0.86, 4.32] | 1.09E-01 | | |

**Supplementary Table 20. Lag sensitivity analyses**. Excluding incident Parkinson's disease events occurring within 2 years of baseline actigraphy assessment. Cox proportional hazards models were refitted after removal of early PD events to evaluate potential reverse causality and subclinical disease effects. Hazard ratios (HRs) from the primary and lagged models are shown for individual prodromal features and cognitive variables, together with 95% confidence intervals (CIs), p-values, analytical sample sizes, and incident PD events. Stability of effect estimates across analyses supports a prodromal rather than reverse-causation interpretation of the observed associations.

| Model | Marker | N | Events | AIC | BIC | Δ AIC | Δ BIC | C-index | Null C-index |
|---|---|---|---|---|---|---|---|---|---|
| M0 RBD categorical | - | 87421 | 448 | 9944.9 | 9973.64 | -89.88 | -81.66 | 0.7823 | 0.7612 |
| M0 RBD continuous | - | | | 9954.76 | 9979.39 | -80.02 | -75.91 | 0.7836 | 0.7612 |
| MA RBD continuous + PRS | - | | | 7706.66 | 7772.68 | -59.33 | -55.44 | 0.8435 | 0.8337 |
| MA RBD categorical + PRS | - | 66524 | 359 | 7701.52 | 7771.42 | -64.47 | -56.7 | 0.8423 | 0.8337 |
| MF RBD × PRS interaction | - | | | 7665.04 | 7742.71 | -193.36 | -173.94 | 0.8422 | 0.7681 |
| M1 Baseline | | | | 3545.6 | 3567.68 | 3.89 | 10.2 | 0.7569 | 0.7567 |
| M2 Additive | Fluid Intelligence | 32668 | 173 | 3508.36 | 3536.74 | -33.35 | -20.74 | 0.7879 | 0.7567 |
| M3 Interaction | | | | 3510.47 | 3551.46 | -31.24 | -6.02 | 0.7872 | 0.7567 |
| M1 Baseline | | | | 10035.6 | 10064.33 | 3.57 | 11.78 | 0.7607 | 0.7613 |
| M2 Additive | Reaction Time | 87185 | 448 | 9945.6 | 9982.54 | -86.43 | -70.01 | 0.7825 | 0.7613 |
| M3 Interaction | | | | 9941.85 | 9995.21 | -90.18 | -57.34 | 0.7817 | 0.7613 |
| M1 Baseline | | | | 5721.52 | 5746.66 | 1.1 | 8.29 | 0.7605 | 0.7563 |
| M2 Additive | Numeric Memory | 50627 | 268 | 5664.6 | 5696.92 | -55.82 | -41.45 | 0.7791 | 0.7563 |
| M3 Interaction | | | | 5667.84 | 5714.52 | -52.58 | -23.85 | 0.777 | 0.7563 |
| M1 Baseline | | | | 10036.43 | 10061.05 | 1.65 | 5.75 | 0.7617 | 0.7612 |
| M2 Additive | Constipation | 87421 | 448 | 9946.61 | 9979.45 | -88.17 | -75.85 | 0.7831 | 0.7612 |
| M3 Interaction | | | | 9949.7 | 9990.75 | -85.08 | -64.55 | 0.7836 | 0.7612 |
| M1 Baseline | | | | 10035.02 | 10059.65 | 0.24 | 4.35 | 0.7613 | 0.7612 |
| M2 Additive | Orthostatic Hypotension | 87421 | 448 | 9945.18 | 9978.02 | -89.6 | -77.28 | 0.7829 | 0.7612 |
| M3 Interaction | | | | 9948.76 | 9989.81 | -86.02 | -65.49 | 0.7829 | 0.7612 |

**Supplementary Table 21. Model fit and discrimination**. Summary across Cox proportional hazards model families. The table reports model performance metrics for baseline prodromal-only models (M1), additive RBD models (M2), interaction models (M3), PRS-adjusted models (MA/MF), and primary RBD-only models (M0). Metrics include Akaike information criterion (AIC), Bayesian information criterion (BIC), partial log-likelihood, concordance statistic (C-index), and comparisons against covariate-only null models. Negative ΔAIC and ΔBIC values indicate improved model fit relative to the null model. Higher C-index values indicate improved discrimination for future Parkinson’s disease incidence prediction.

### Figures

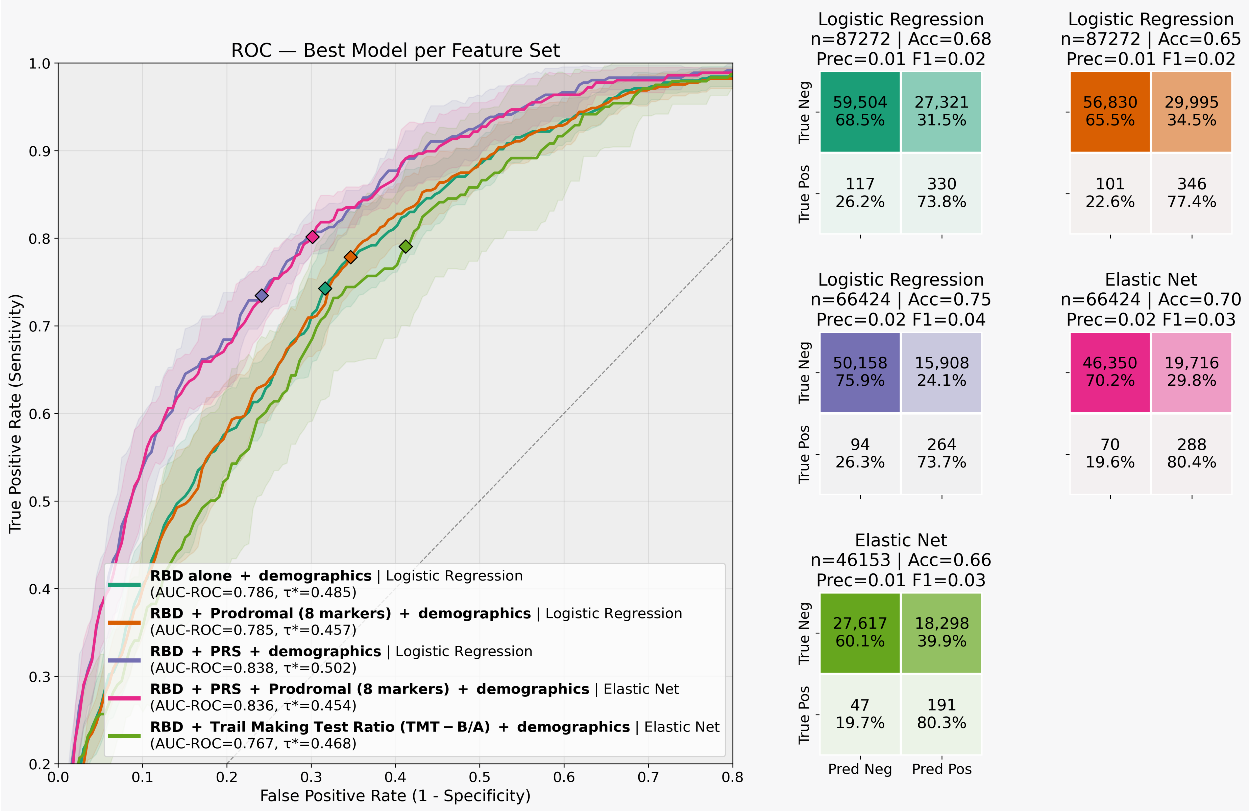


**Supplementary Figure 1. Comparative performance of cross-sectional machine learning models for identifying future incident Parkinson's disease cases using accelerometry-derived RBD risk, prodromal markers, genetic risk, and cognitive features.** Receiver operating characteristic (ROC) curves are shown for the best-performing model within each feature-set configuration. All models included demographic covariates (age, sex, and body mass index), with additional predictors consisting of accelerometry-derived RBD probability scores, prodromal markers, Parkinson's disease polygenic risk scores (PRS), and the Trail Making Test ratio (TMT-B/A). Model performance was evaluated using nested cross-validation and restricted to incident Parkinson's disease cases in the test folds.
